# CAR^+^ and CAR^-^ T cells differentiate into an NK-like subset that is associated with increased inflammatory cytokines following infusion

**DOI:** 10.1101/2022.03.29.22273013

**Authors:** Raymond Hall Yip Louie, Curtis Cai, Mandeep Singh, Ira Deveson, James Ferguson, Timothy G. Amos, Helen Marie McGuire, Jerome Samir, Kavitha Gowrishankar, Thiruni Adikari, Robert Balderas, David Bishop, David Gottlieb, Emily Blyth, Kenneth Micklethwaite, Fabio Luciani

**Affiliations:** Kirby Institute for Infection and Immunity, UNSW Sydney, Australia; School of Medical Sciences, UNSW Sydney, Australia; Garvan Institute for Medical Research, Sydney, Australia; Ramaciotti Facility for Human Systems Biology, The University of Sydney, Sydney, NSW, Australia. Charles Perkins Centre, The University of Sydney, Sydney, NSW, Australia; Infection, Immunity and Inflammation Theme, School of Medical Sciences, Faculty of Medicine and Health, The University of Sydney, Camperdown, NSW, Australia; Blood Transplant and Cell Therapies Program, Department of Haematology, Westmead Hospital, Sydney, NSW, Australia; Westmead Institute for Medical Research, Sydney, NSW, Australia; Becton Dickinson, San Jose, CA, USA; Sydney Medical School, The University of Sydney, Sydney, NSW, Australia; NSW Health Pathology Blood Transplant and Cell Therapies Laboratory – ICPMR Westmead, Sydney, NSW, Australia

**Author notes:** Corresponding author: Associate Professor Fabio Luciani Immunogenomics laboratory, School of Medical Sciences, and the Kirby Institute University of New South Wales Sydney 2052, Australia.

**Keywords:** CAR T cells, NK-like, single cell multi-omics, trajectories, immune-depletion, clonal expansion, immune-reconstitution

## Abstract

Chimeric antigen receptor (CAR) T cells have demonstrable efficacy in treating B-cell malignancies. Factors such as product composition, lymphodepletion and immune reconstitution are known to influence functional persistence of CAR^+^ T cells. However, little is known about the determinants of differentiation and phenotypic plasticity of CAR^+^ T and immune cells early post-infusion. We report single cell multi-omics analysis of molecular, clonal, and phenotypic profiles of CAR^+^ T and other immune cells circulating in patients receiving donor-derived products. We used these data to reconstruct a differentiation trajectory, which explained the observed phenotypic plasticity and identified cell fate of CAR^+^ and CAR^-^ T cells. Following lympho-depletion, endogenous CAR^-^ CD8^+^ and γ□ T cells, clonally expand, and differentiate across heterogenous phenotypes, from a dominant resting or proliferating state into precursor of exhausted T cells, and notably into a terminal NK-like phenotype. In parallel, following infusion, CAR^+^ T cells undergo a similar differentiation trajectory, showing increased proliferation, metabolic activity and exhaustion when compared to circulating CAR^-^ T cells. The subset of NK-like CAR^+^ T cells was associated with increasing levels of circulating proinflammatory cytokines, including innate-like IL-12 and IL-18. These results demonstrate that differentiation and phenotype of CAR^+^ T cells are determined by non-CAR induced signals that are shared with endogenous T cells, and condition the patients’ immune-recovery.

**One Sentence Summary:** CAR^+^ and CAR^-^ CD8+ T cells share a differentiation trajectory terminating in an NK-like phenotype that is associated with increased inflammatory cytokines levels.

**Graphical Abstract:** 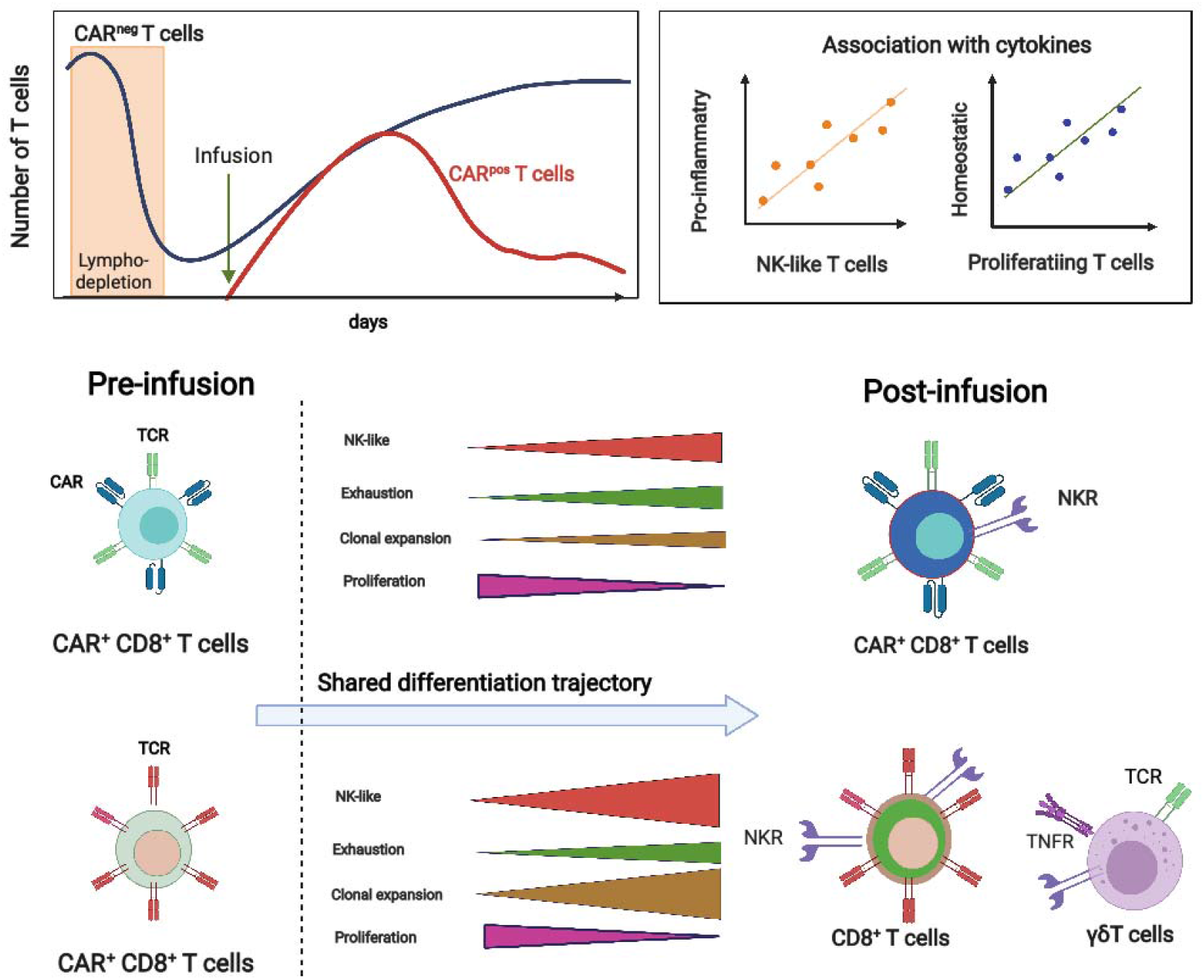

## INTRODUCTION

Chimeric antigen receptor (CAR) T cells targeting CD19 are used for the treatment of relapsed haematological malignancies such as diffuse large B cell lymphoma (DLBCL) and B cell acute lymphoblastic leukaemia (B-ALL)^1–3^. There are many factors influencing the success of these therapies, such as cytokine release syndrome (CRS), neurotoxicity, infection due to pre-infusion lymphodepleting chemotherapy, immune reconstitution, relapse owing to loss of the CD19 epitope in the tumor, and persistence of functional CAR T cells^4, 5^. For example, previous work characterizing the phenotype of CAR T cells in the infusion product (IP) showed that poor response was correlated with an exhaustion phenotype^6^, while remission at 3-month follow-up had three-fold higher frequencies of endogenous CD8^+^ T cells expressing central memory signatures^7^. Immuno-conditioning with cyclophosphamide/fludarabine prior to cell infusion has been also associated with CAR T cell persistence in patients^8, 9^. The kinetics of CAR T cells in peripheral blood typically peaked at 10 to 14 days post-infusion and then declined slowly over time. Complete response has been associated with higher levels of circulating CAR T cells, and their expansion correlated with severity of cytokine release syndrome (CRS)^10^. However, there is limited knowledge regarding the cellular and molecular evolution of CAR T cells following infusion, which may impact therapy success. This requires an understanding of the molecular and phenotypic composition of not only the IP, but the endogenous immune reconstitution post-infusion, as this may affect CAR T cell kinetics, for instance via clonal expansion of T cells recognising pathogens, or immunological rejection of the CAR T cells^7–9^.

Single-cell RNA seq (scRNA-seq) and clonal analysis was recently used to investigate the phenotypic profile and evolution of autologous CD19 CAR T cells in the product^7, 11, 12^ and circulating at post-infusion^13^, and in one patient treated with B cell maturation antigen (BMCA) CAR T cells^14^. These studies showed similar trends of CAR T cells comprising both a proliferative and cytotoxic phenotype, which were recently recapitulated using *in vitro* cell culture models and single-cell multi-omics revealing distinct functional and phenotypic profiles following *in vitro* stimulation^12^. The latter also revealed molecular differences in cell composition between patient- and donor-derived products. A recent *in vitro* model for mesothelin-directed CAR against pancreatic cancer showed that following persisting antigen stimulation, CAR T cells develop a dysfunctional state with an NK-like phenotype, which was regulated by transcription factors ID3 and SOX4^15^.

While these findings provide some understanding of how CAR T cell differentiation and plasticity drive the evolution of infused cells, more investigation is required to understand these processes *in vivo*, and how the recovering immune system may interact with CAR T cells and influence their differentiation and function. For example, despite HLA-matched conditions, antigen stimuli may interfere with CAR T cell functions via minor HLA-molecules or other mechanisms involving NK and *γ*δ T cells^16^. These unconventional immune cells are known to play a key role in controlling infections^17^, interfering with the tumor micro-environment^18^, and regulating allogenic responses following hematopoietic stem cell transplant (HSCT)^19^. The study of CAR-immune dynamics is important in the context of donor-derived CAR T cells, as several trials are underway utilising donor-derived products and there is limited understanding of the associated advantages and risks^20^, such as CAR T cell derived lymphoma as recently reported by our group^21, 22^.

In this study we performed a single-cell multi-omics analysis with simultaneous measures of scRNA-seq, protein expression profiles and T cell receptor (TCR) sequences, as well as mass-cytometry investigation of patients’ pre- and post-infusion circulating immune cells, as well as CAR^+^ T cells from IP and post-infusion samples from a donor-derived CAR19 T cell clinical trial^21^ (Fig. 1A). This analysis revealed a common differentiation trajectory shared by CAR^+^, as well as both endogenous conventional CAR^-^ CD8^+^ and unconventional γδ T cells. While some of these findings confirm the reported phenotypes acquired by CAR T cells following infusion^13, 14^, here we discovered similarities in the cell fate and phenotypic plasticity observed in the endogenous subsets of CD8^+^ and γδ T cell subsets, and the donor-derived CAR T cells. We provide evidence that TCR-signalling and homeostatic proliferation contribute to the differentiation and phenotype of CAR T cells, and finally reveal associations between serum cytokines and molecular signatures characterising subsets of both CAR^+^ and CAR^-^ T cells. This comprehensive molecular, cellular and clonality atlas provides an explanation for the observed dynamics of CAR T cells, and their interaction with the reconstituting immune cells following lymphodepletion.

**Fig. 1.**
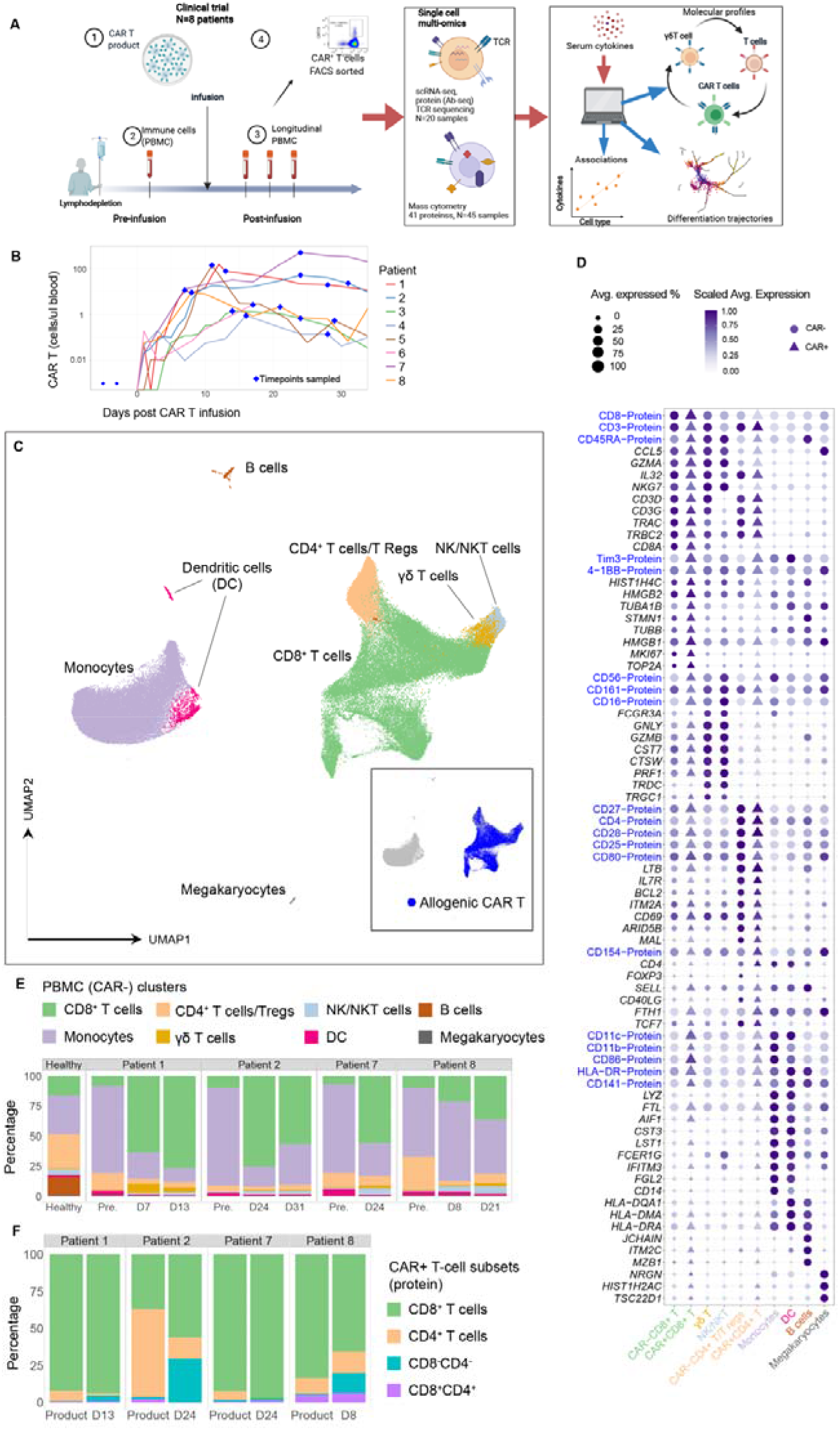
Longitudinal analysis of the molecular and cellular composition of allogenic CAR T and endogenous patient immune cells from pre- to post-infusion. **(A)** Overview of study design. For each patient (N=8), peripheral blood mononuclear cells (PBMC) from pre- and post-infusion samples, and CAR T cells from product and post-infusion samples were analysed. Single cell multi-omics (N=4) and mass cytometry (N=8) were utilised to generate molecular profiles of immune cells, and these were then used to identify differentiation trajectories and associations with cytokines. **(B)** Kinetics of CAR T cells following infusion, blue symbols indicate sample time points utilised for downstream multi-omics. **(C)** Dimensionality reduction using UMAP of CAR^+^ and CAR^-^ T cells using single cell transcriptomics showing canonical cell types. **(D)** Dot plot of selected proteins and genes. The size of each point represents the percentage of cells with non-zero expression. Colours represent the scaled log expression averaged over all endogenous or CAR T samples. CAR^-^ cells are represented by filled circles, CAR^+^ by filled triangles. (**E)** Distribution of canonical cell types in the CAR^-^ samples. **(F)** Distribution of CD8^+^ and CD4^+^ subsets in the CAR^+^ samples.

## RESULTS

### Single-cell atlas of PBMC and CAR T cells reveals post-infusion expansion of CD8^+^ T cells

We investigated eight patients from a clinical trial^21^ (Supplementary Table S1) with relapsed or refractory B cell malignancy following HSCT who were subsequently treated with second generation CAR T cells (Methods). Seven of these patients (P1-P5, P7, P8) received IP generated from cells obtained from the same sibling HSCT donor, and one patient (A1) had an autologous CAR T transfer expressing an identical CAR to the allogeneic (donor-derived) IP. All patients had engrafted before infusion of CAR T cells except for P3, who experienced partial donor marrow chimerism at the time of administration of CAR T and thus the endogenous immune repertoire was not allogenic to the CAR T cells. Patients underwent cyclophosphamide/fludarabine lymphodepletion conditioning for three days prior to infusion, resulting in cytopenia (Fig. 1A). Robust expansion of CAR T cells was observed with peak count occurring within the first 2 weeks (Fig. 1B). As previously reported, two patients (P2 and P8) developed CAR T cell derived lymphoma^21, 22^.

We sought to understand the evolution of the reconstituting immune populations, and of the allogenic CAR T cells during the initial stage of therapy (Fig.1A). Single cell multi-omics data were thus generated from peripheral blood mononuclear cells (PBMC) one day prior to CAR T cell infusion and up to a month post-infusion. CAR T cells were obtained from the IP and from flow cytometric sorting of CD3^+^CAR^+^ cells from post-infusion PBMC samples (Fig. 1A, Extended Data Fig. 1A). A total of 106,976 cells were analysed from four patients (P1, P2, P7 and P8), comprising of single cell measures of gene and extracellular protein expression, as well as full length T cell receptor (TCR) sequences across 20 samples including 11 PBMC and 8 CAR T cell sorted populations and PBMCs from one healthy donor as a reference population (Supplementary Tables S2 and S3). Mass cytometry (CyTOF) was also utilised to longitudinally characterise protein expression profiles of 41 extracellular and intracellular protein markers for PBMCs and CAR T cells obtained from 45 samples of 8 patients (Supplementary Table S4, Fig. 1A). We also identified CAR^+^ T cells from the scRNA-seq data by detecting CAR T transcripts (Methods), which showed a positive correlation with the number of circulating CAR^+^ T cells identified using an anti-CAR antibody in the PBMC samples (Extended Data Fig. 1B). Circulating CAR T cells were at low frequencies within the PBMC populations, hence we will refer to CAR^+^ cells as those isolated via flow cytometry using the anti-CAR antibody, and CAR^-^ cells as cells identified from the PBMC samples.

Dimensionality reduction and clustering of gene and protein expression profiles revealed eight clusters identifying canonical cell types (Fig. 1, C and D), with gene and protein expression profiles consistent between patients (Extended Data Fig. 1C, Supplementary Table S5). These comprised of CD8^+^ T cells, CD4^+^ T/regulatory T CD25^+^*FOXP3*^+^ cells, NK/NKT, *γ*δ T cells, B cells, dendritic cells (DCs), megakaryocytes, and monocytes (Fig. 1C). The population of monocytes decreased in post-infusion samples across all patients (Fig. 1E). As expected, only a small number of B cells were identified in the blood.

The largest cluster was formed by CD8^+^ T cells including both CAR^-^ (Fig. 1E) and CAR^+^ (Fig. 1F) cells, which was consistent with the CyTOF data for all eight patients (Extended Data Fig. 1, D and E). Both CAR^+^ and CAR^-^ CD8^+^ T cells expressed intermediate levels of cytotoxic (*GZMA*, *PRF1*, *IL32*) and NK-like markers (CD56, CD16, CD161, *NKG7, FCGRA*) (Fig. 1D, Extended Data Fig. 1F), suggesting that both the CAR^-^ and CAR^+^ CD8^+^ T population comprised of cytotoxic NK-like (innate-like) CD8^+^ T cells^23–25^. All patients demonstrated an increase in the proportion of CAR^-^ CD8^+^ T cells (Fig. 1E, Extended Data fig. 1, G and H), and similar increases were found for γδ T cells (Fig. 1E). The majority of CAR^+^ T cells were found in the CD8^+^ subset except for the IP in patients P2, P4 and A1 which were dominated by CD4^+^ (Fig. 1F, Extended Data fig. 1E). Following infusion, CAR^+^ T cells increased in the CD8^+^ and CD4^-^CD8^-^ subsets (Fig. 1F, Extended Data fig 1E). In summary, single-cell multi-omics and mass cytometry revealed significant expansion of CD8*^+^* and γδ T cells following infusion, with CAR^+^ and CAR^-^ T cells sharing similar molecular profiles.

### CAR^-^ and CAR^+^ T cells form functionally distinct subsets including an NK-like phenotype

To further explore the heterogeneity of the CD8*^+^* T cell population we performed unsupervised analysis within the subset of cells identified as CD3^+^CD4^-^CD8^+^, which comprised of both CAR^-^ and CAR^+^ T cells. After removing a cluster with high levels of CD56 residing in the NK/NKT subset (Fig. 1C), we identified 6 clusters (C1-C6) (Fig. 2A) each including product, pre- and post-infusion CAR^-^ and CAR^+^ T cells (Fig. 2, B and C, Extended Data fig. 2A).

**Fig. 2.**
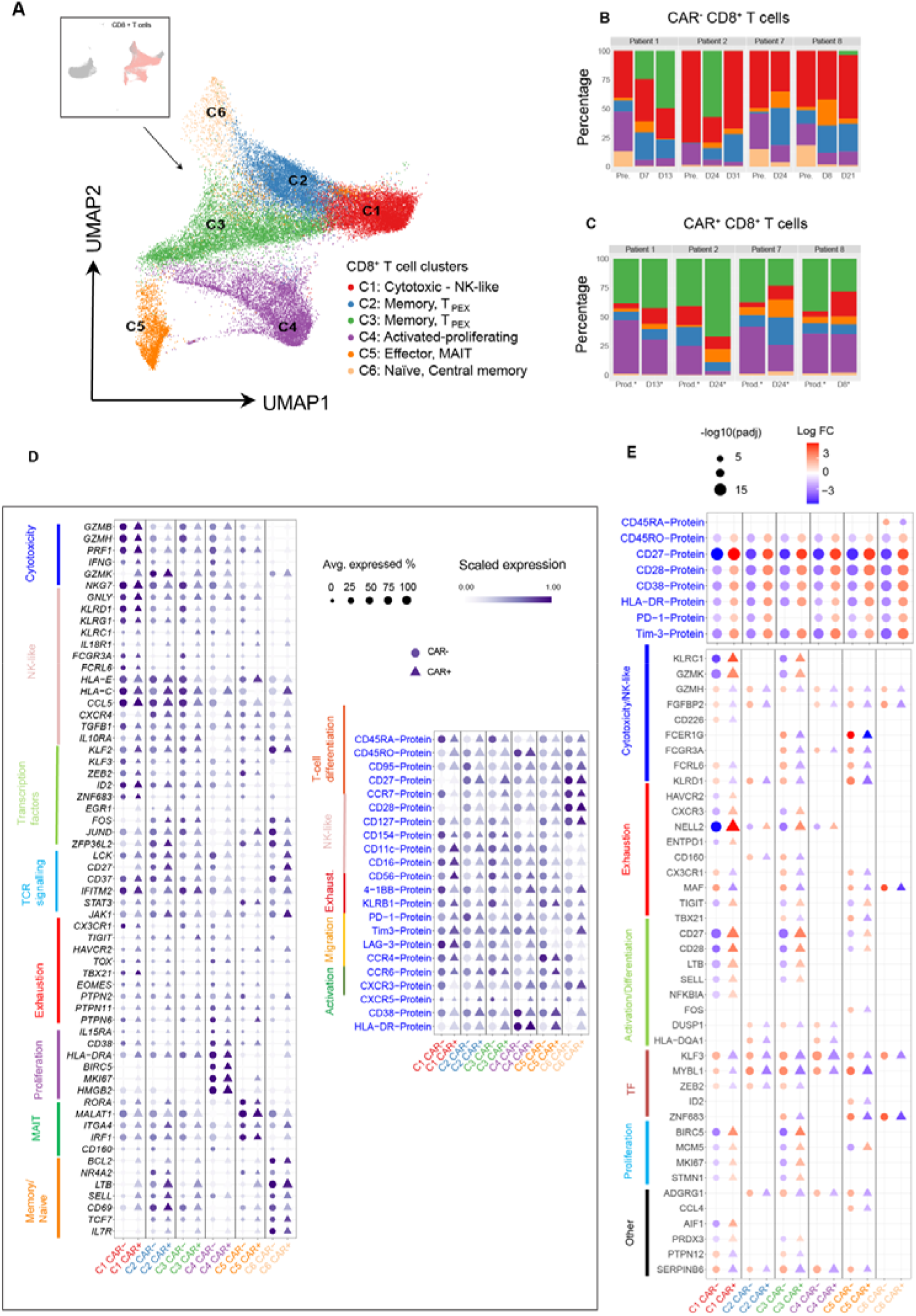
Single-cell multi-omics identify heterogenous subsets of CD8^+^ T cells shared between CAR^-^ and CAR^+^ T cells. **(A)** Dimensionality reduction (UMAP) of CD8*^+^* T cells from both CAR^+^ and CAR^-^ samples using scRNA-seq data, revealing six clusters. **(B-C)** Distribution of the clusters by sample time-point and patient for both CAR^-^ (B) and CAR^+^ T cells (C). **(D)** Dot plots of expression values for selected genes and proteins (AbSeq). Shown are gene markers with significant p-value (<0.1 using MAST). The size of each point represents the percentage of cells with non-zero expression. The colour represents the scaled log expression averaged over all CAR^-^ or CAR^+^ T samples. CAR^-^ samples are represented by filled circles, CAR^+^ T samples by filled triangles. **(E)** Dot plot of fold change values for gene and proteins that were differentially expressed between CAR^+^ and CAR^-^ CD8^+^ T cells circulating in post-infusion samples. Differential expression analysis was performed using patients as replicates. The size of each point represents log10(adjusted p-value). The colour represents the log fold change. CAR^-^ T cells are represented by filled circles, CAR^+^ T samples by filled triangles.

Differential gene and protein expression analysis (Supplementary Table S5, Fig. 2D) was then performed to identify cluster phenotypes. Cells in C6 comprised of resting cells and included both naïve and central memory T cells expressing CD27 and CD127 (Fig. 2D), and transcription factors associated to resting state, such as *TCF7, IL7R*, *SELL* (Fig. 2D). Cluster C5 revealed an effector memory profile, and proteins associated with tissue homing (CCR4, CCR6), and unconventional mucosal associated invariant T (MAIT) cells expressing high levels of KLRB1 (CD161), and genes *IL18R1*, *CXCR4*, *MAF*, *SLC4*^24, 26–28^. Cells within C4 expressed activation protein markers CD38 and HLA-DR, and genes associated to proliferation, e.g., *MKI67, HMGB2, BIRC5*, as well as homeostatic cytokine receptors *IL15RA*. Cells in C4 expressed exhaustion proteins Tim-3, PD-1, and genes *TOX*, *TIGIT*, *CX3CR1*^6, 28, 29^, as well as Protein Tyrosine Phosphatase Non-Receptors *PTPN11* (encoding SHP-2) and *PTPN6* which have been associated with exhaustion^30, 31^ (Fig. 2D). This profile, together with a distinct cell-cycle signature (Extended Data Fig. 2B), is consistent with the highly proliferative dysfunctional subset identified within tumour-infiltrating CD8^+^ T cells in melanoma^29^. Clusters C2 and C3 comprised of cells with both effector memory phenotype, expressing activation markers CD38 and HLA-DR, and CD27, with C3 largely formed by CAR^+^ T cells (Fig. 2C, Extended Data fig. 2A). These two clusters revealed progenitor of exhausted cells (T_PEX_), having intermediate levels of proteins PD-1, Tim-3 and LAG-3, (Fig. 2E) and genes *TIGIT*, *GZMK*, *TOX, PTPN1* and *PTPN11 (SHP2)*^32^ (Fig. 2D). These cells retained levels of AP-1 transcription factors *FOS* and *JUN*, which are known to be associated with resistance to exhaustion in CAR T cells^33^.

We discovered a shared NK-like signature among CAR^+^ and CAR^-^ cells within cluster C1. C1 expressed predominantly a T_EMRA_ phenotype (CD45RA^+^CD27^-^CD28^-^) and expressed NK-like markers CD56, CD16, CD11c, and KLRB1 (Fig. 2D) at comparable levels to those expressed by NK and NKT cells (Extended Data Fig. 1F). This NK-like profile in CD8^+^ T cells has been previously observed in viral infection^34^ and cancer^23^, as well as in CAR T cells under persisting stimulation *in vitro*^15^. These cells also expressed CD154 (and gene *CD40L*) which is known to induce T cell expansion via interactions with dendritic cells producing IL-12p70^35^, as well as *IL10RA* and *TGFB*, which are involved in the inhibition of cytotoxicity in NK cells^36^. Besides expression of classical CD8 cytotoxicity marker genes (e.g., *PRF1*, *IFNG*, *GZMB*) (Fig. 2D), these cells expressed *NKG7, KLRG1, KLRD1*, and *IL18R1* (Fig. 2D), and transcription factors *KLF2*, *ZEB2*, and *TBX21* which are involved in regulation of T cell differentiation and trafficking^37–39^. The NK-like profile was also detected in the T_PEX_ subsets (C2 and C3), although the expression of genes and proteins characterising this signature was reduced compared to C1 (Fig. 2D). These three clusters (C1-C3) shared signatures of activation (*LCK*) and homing (*CCL5*), as well as high levels of *HLA-E*, and *HLA-C*, which are ligands for NKG2A and KIR, respectively, which indicate interactions with NK cells.

Gene Set Enrichment Analysis (GSEA) confirmed the distinct gene signatures of each cluster, notably the enrichment of both CD8 T cell cytotoxicity as well as NK-like signature in clusters C1 (Extended Data Fig. 2C, Supplementary Table S6). Cells in clusters C1, C2 and C3 had enriched exhaustion profile in line with in vitro findings^15^.

We next sought to identify phenotypic changes of CAR^+^ cells as they molecularly and phenotypically evolve from product to circulating post-infusion, and CAR^-^ cells from pre- to post-infusion. The key finding is that both CAR^-^ and CAR^+^ CD8^+^ T cells had an up-regulation of NK-like markers at post-infusion, suggesting evolution towards an NK-like profile. Specifically, focusing first on CAR^+^ cells, we found circulating CAR^+^CD8^+^ T cells had increased cytotoxic, exhausted and NK-like profiles, and decreased memory-like phenotype compared to the infusion product (Extended Data Fig. 3, A and B). Activation (CD27, CD28), inhibitory (PD-1), and NK-like (KLRB1 (CD161)) markers were up-regulated in circulating cells compared to product, across all clusters (Extended Data Fig. 3A). Gene expression revealed extensive differences (26, 971 differential expressed genes, adjusted p-value < 0.1, Extended Data Fig. 3B, Supplementary Table S5) which confirmed circulating cells had increased cytotoxic (*GZMH*), NK-like signatures (*IFNG, KLRC1, KLRG1, FCRL6, CD160)*, TCR signalling (*ZAP70*), and T_PEX_ markers (*TIGIT*, *LAG-3*, *TOX,* and *CX3CR1*)^32, 40^. Circulating cells also had increased levels of transcription factors associated with regulation of T cell differentiation such as *BATF*^41, 42^, and *IRF9* and *TGFB* which are known to regulate T cell effector functions^43, 44^ (Extended Data Fig. 3B). In contrast, cells in the IP expressed higher levels of memory markers *IL7R*, *IL2RA* as well as transcription factors controlling T cell activation and favouring memory formation, *EGR1*, *BATF3*^41^, and *BACH2*^42^. CAR^-^ CD8^+^ T cells had significantly less differentiated genes from pre- to post-infusion across all clusters (424 genes, adjusted p-value <0.1, Extended Data Fig. 3C, Supplementary Table S5), but showed similar increases in cytotoxic (*GZMB, GZMH*) and NK-like markers (*FCGR3A* (CD16), *NKG7*) from pre- to post-infusion (Extended Data Fig. 3C).

Finally, we validated the heterogenous CD8^+^ T clusters including the NK-like subset across the 8 patients, utilising mass cytometry data (Extended Data Fig. 4, A-E). These data confirmed the presence of phenotypically distinct subsets of CD8^+^ T cells and four major clusters (M1-M4) classified into cytotoxic M1 (KLRG1, Perforin, Granzyme B, T-bet), memory T_PEX_ M2 (CD69, CXCR3, CD39), naïve and central memory M4 (SELL, IL7R, CD28, CD45RA), and finally a proliferation/activation subset M3 (CD38, HLA-DR, KI67). Notably, the NK-like signature we previously observed in cytotoxic C1 and to a less extent within the memory T_PEX_ clusters (C2 and C3) was confirmed via mass cytometry in all patients, with a high expression of CD56, CD16, and CD49D^23, 24^ in the cytotoxic (M1) and to a less extent in the memory (M2) clusters (Extended Data Fig. 4, C and E).

In summary, our findings from both multi-omics and mass cytometry identified the presence of cytotoxic, exhausted, NK-like CD8^+^ T cells which are present in both CAR^-^ and CAR^+^ subsets.

### Circulating CAR^+^ CD8^+^ T cells differ from CAR^-^ CD8^+^ T cells by increased activation and proliferation with distinct NK-like profile

Focusing on post-infusion samples, we reasoned that despite the similarities, CAR^+^ and CAR^-^ CD8^+^ T cells also differ in molecular and phenotypic markers within each cluster. We observed that CAR^+^ T cells had a higher level of surface markers consistent with increased activation (CD27, CD38 and HLA-DR) and exhaustion (Tim3, PD-1) across all clusters (Fig. 2E). Transcriptionally, we observed even greater distinction between CAR^+^ and CAR^-^ T cells within clusters. Notably, CAR^+^ T cells within cytotoxic (C1) and memory T_PEX_ (C3) clusters showed up-regulation of cytotoxic (*GZMK, FGFBP2*), NK-like (*KLRC1* encoding NKG2A) and exhaustion markers (*TIGIT*, *CXCR3*, *NELL2, PRDX3*)^27, 28^ (Fig. 2G, Supplementary Table S5). In contrast, CAR^-^ T cells up-regulated cytotoxic gene *GMZH,* chemokine *CXCR3,* NK-like marker *KLRD1* (*CD94*), and transcription factors *MYBL1* and *KLF3*, and ZEB2 which is known to drive T cell differentiation^39^. CAR^-^ T cells in memory T_PEX_ C3 and effector C5 also had up-regulation of transcription factors *TBX21* and *ZNF683* (Hobit), and Fc-gamma receptors (*FCGR3A*, *FCER1G,* and *FCRL6)*. GSEA also identified post-infusion CAR^+^ T cells were enriched in pathways for metabolic activities (oxidative phosphorylation, electron transport), DNA replication but a reduced enrichment in regulatory/inhibitory functions mediated by TNF and NK receptors (Supplementary Table S6), compared to CAR^-^ cells. Taken together, this suggests that although post-infusion CAR^-^ and CAR^+^ CD8^+^ T cells share an up-regulated cytotoxic-NK like phenotype compared to the product and pre-infusion samples, there are specific cytotoxic and NK-like markers which delineate CAR^-^ and CAR^+^ cells at post-infusion. Other distinguishing cluster specific differences were also observed, related to differentiation, and regulation of activation and proliferation.

### CAR^-^ and CAR^+^ CD8^+^ T cells shared a differentiation trajectory through T_PEX_ towards cytotoxic and NK-like state

We investigated the extent of clonal expansion of both CAR^-^ and CAR^+^ CD8^+^ T cells in the previously identified CD8^+^ T phenotypes, reasoning that the observed shared distribution of cell states reflects the presence of common mechanisms underlying the differentiation process of both subsets. In each subset, clonal expansion was quantified by identifying full-length αβ-TCR sequences from the single cell data utilising long-read sequencing^45^. A total of 2054 and 1016 clones were identified in CAR^-^CD8^+^ and CAR^+^CD8^+^ T cells, respectively (Extended Data Fig. 5, A and E). Focusing first on CAR^-^CD8^+^ T cells, we observed an increase in the number of expanded clones from pre- to post-infusion (Fig. 3A), and the Shannon evenness index, which is a measure of the degree of clonal diversity, revealed a reduction in diversity in post-infusion samples (Extended Data Fig. 5B). Clonal expansion in post-infusion samples occurred mainly in cytotoxic (C1) and memory T_PEX_ clusters (C2, C3) (Fig. 3B), which contrasted with the distribution of clones identified in pre-infusion samples, which were expanded in the cytotoxic (C1) and the proliferative (C4) subsets (Extended Data Fig. 5C). The majority of clones that were detected in pre-infusion samples persisted following the infusion of CAR T cells across all clusters, with the majority of them mapped within the cytotoxic cluster (Fig. 3C). Persisting clones showed transition between clusters from pre- to post-infusion samples, thus confirming intra-clonal phenotypic plasticity (Extended Data Fig. 5D).

**Fig. 3.**
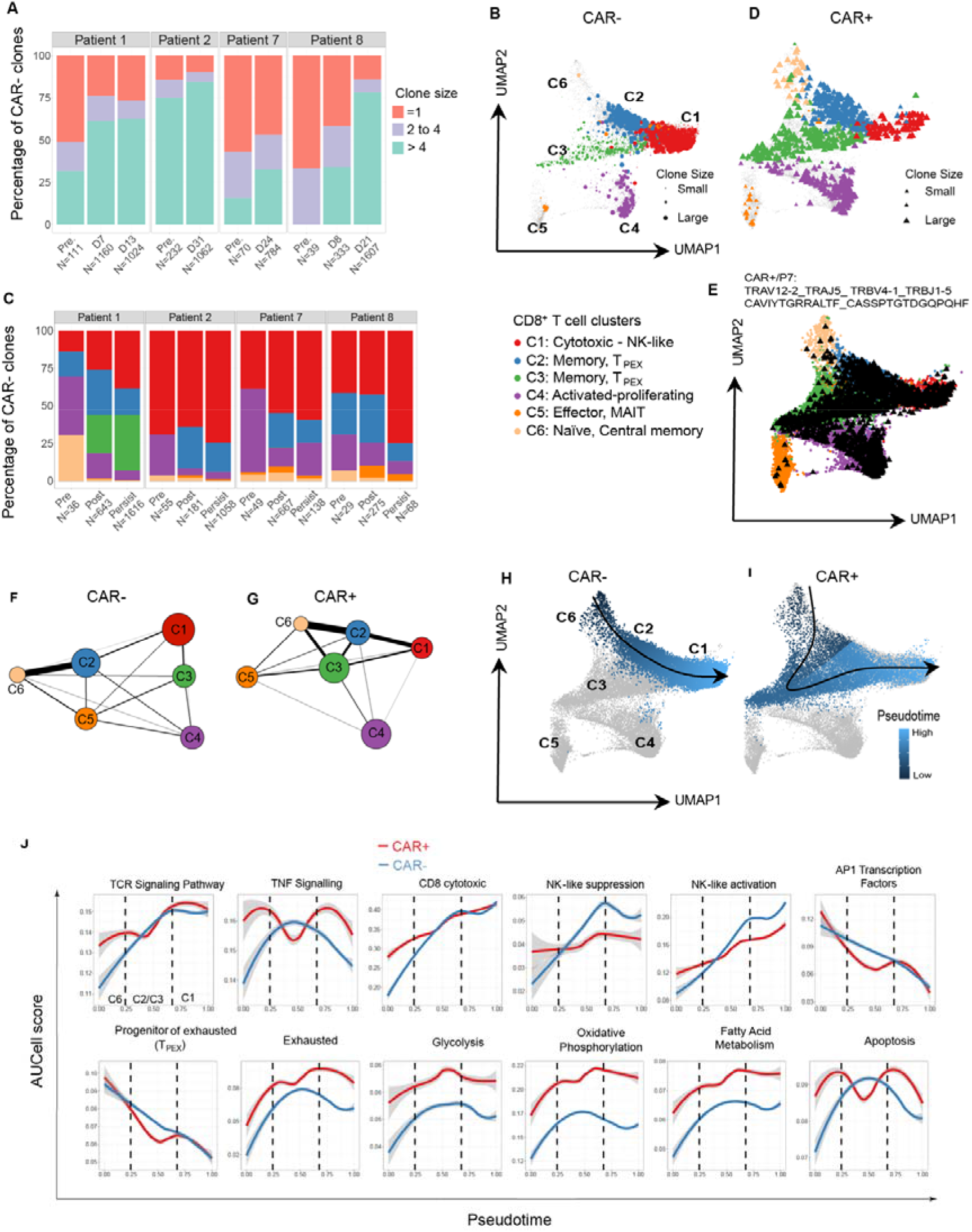
Clonal and trajectory analyses reveal differentiation and fate of CAR^-^ and CAR^+^ CD8^+^ T cells during therapy. (**A**) Distribution of clone size identified in the endogenous CAR^-^CD8^+^ T cells. (**B**) Post-infusion expanded clones (size > 1) identified in CAR^-^ cells, mapped on the UMAP plot. **(C)** Proportions of clones that were unique to pre- or post-infusion samples, or were persisting from pre- to post-infusion. (**D**) Post-infusion expanded clones (size > 1) identified in CAR^+^ cells, mapped on the UMAP plot. (**E**) Mapping of a single clone with a TCRαβ that was matched to a CMV specific TCR sequence identified in the CAR^+^ T cells in P7, showing heterogenous distribution across all clusters. (**F-G**) PAGA graph obtained from CAR^-^ (F) and CAR^+^ CD8^+^ T cells (G), revealing cluster connectivity (represented by line weights). (**H-I**) Pseudotime values along inferred differentiation trajectory for CAR^-^ (H) and CAR^+^ CD8^+^ T cells (I). **(J)** Loess curves of gene modules calculated by AUCell versus the scaled pseudotime values along the inferred trajectory utilising only cells from post-infusion samples. Gray shade represents 95% confidence interval.

CAR^+^CD8^+^ T cells were characterised by a more diverse clonal repertoire with reduced clonal expansion when compared to the endogenous subset (Extended Data Fig. 5F). CAR^+^CD8^+^ clones were found across all clusters, predominantly within the proliferating (C4) and T_PEX_ (C3) clusters (Fig. 3D, Extended Data fig. 5G), largely reflecting the higher proportion of CAR^+^ T cells within these two clusters. TCR clonal analysis of two IPs which were administered in P2 and P8 revealed a very diverse repertoire with no evidence of large clones, thus suggesting a highly diverse TCR repertoire in the IP.

We interrogated the VDJdb database of TCR sequences with known epitope specificity^46^ to identify epitope-specificity of these receptors. For both CAR^-^ and CAR^+^ T cells, we identified TCRs with specificity for influenza, CMV and EBV epitopes (Supplementary Table S7). An exception was P7, where a single clone comprised 61% of the circulating CAR^+^ cells (Fig. 3E). This clone was identified in each of the 6 clusters and carried a TCR αβ that matched a known sequence specific for human cytomegalovirus (CMV) epitope (Supplementary Table S7). Collectively, this data suggests that both CAR^-^ and CAR^+^ T cells undergo clonal expansion and displayed intra-clonal phenotypic plasticity, with evidence of clonal expansion of pathogen specific T cells.

We next set to investigate the dynamics and differentiation trajectory between cell clusters, and identify the relationship between the observed cell states. For this purpose, we applied established bioinformatic tools on the gene expression data, by first constructing a graph depicting inter-cluster connectivity using PAGA^47^ (Fig. 3, F and G). There was a strong predicted connectivity between C6, C2 and C1 for CAR^-^ T cells (Fig. 3F), and between clusters C6, C2, C3, and C1 for CAR^+^ T cells (Fig. 3G). To identify the directional transitions between these clusters and hence construct the differentiation trajectory, we applied RNA velocity^48^, which confirmed the inferred connectivity between clusters and identified a differentiation process from naïve and central memory C6 into an intermediate state identified as the memory T_PEX_ cluster C2 (CAR^-^) or C2 and C3 (CAR^+^), and finally towards the terminally differentiated cytotoxic NK-like state (C1) (Extended Data Fig. 6, A and B). These two trajectories differed only in the addition of the intermediate cluster C3 in the CAR^+^ trajectory, consistent with the prevalence of this cluster in CAR^+^ T cells (Extended Data Fig. 2A). Further evidence of these trajectories was provided by inference of directional transition probabilities between clusters utilising CellRank^25^, which revealed a shared cluster transition pattern for both CAR^-^ (Extended Data Fig. 6C) and CAR^+^ T subsets (Extended Data Fig. 6D). Both RNA velocity and CellRank also revealed that cells within the C4 and C5 clusters were characterised by a self-cycling state (Extended Data Fig. 6, A-D).

We calculated pseudotime values associated with the CAR^-^ (Fig. 3H) and CAR^+^ (Fig. 3I) trajectories to quantify the differentiation and fate from the root cluster C6 to the terminal state C1. Along these two trajectories, we quantified the changes in gene signature scores (Fig. 3J, Extended Data fig. 6E) which revealed increasing TCR signalling, cytotoxicity and NK-like activation signatures, consistent with an increased clonal expansion into the terminal state (C1) (Fig. 3J). In contrast, declining trends were detected for T_PEX_ cells and AP-1 transcription factors. CAR^-^ T cells had an increased NK-like suppression signature compared with CAR^+^ cells but decreased metabolic signatures, e.g., oxidative phosphorylation and glycolysis, and MTOR signalling (Fig. 3J, Extended Data fig. 6E). These profiles were confirmed by protein expression profiles along the trajectory (Extended Data Fig. 6F), showing increased expression of CD45RA and CD56, confirming the increase of T_EMRA_ and NK-like signatures.

In summary, phenotypic and clonal data are consistent with a shared trajectory between CAR^-^ and CAR^+^ T cells which terminated at a cytotoxic NK-like phenotype and identified a TCR-dependent contribution to the evolution of CD8^+^ T cells.

### Clonally expanded **γδ** T cells revealed phenotypic plasticity and repertoire diversity

We next analyzed transcriptional and protein profiles of γδ T cells under the hypothesis that the heterogeneous molecular and phenotypic subsets form a differentiation trajectory similar to that followed by CD8^+^ T cells. Gene expression profile of γδ T cells, identified as CD3^+^CD4^-^CD8^-^ *TRDC*^+^ subset (Extended Data Fig. 7A), revealed 7 clusters (G0-G6) (Fig. 4A), which confirmed a similar molecular profile with CD8^+^ T cells (Fig. 4B, fig. 2D).

**Fig. 4.**
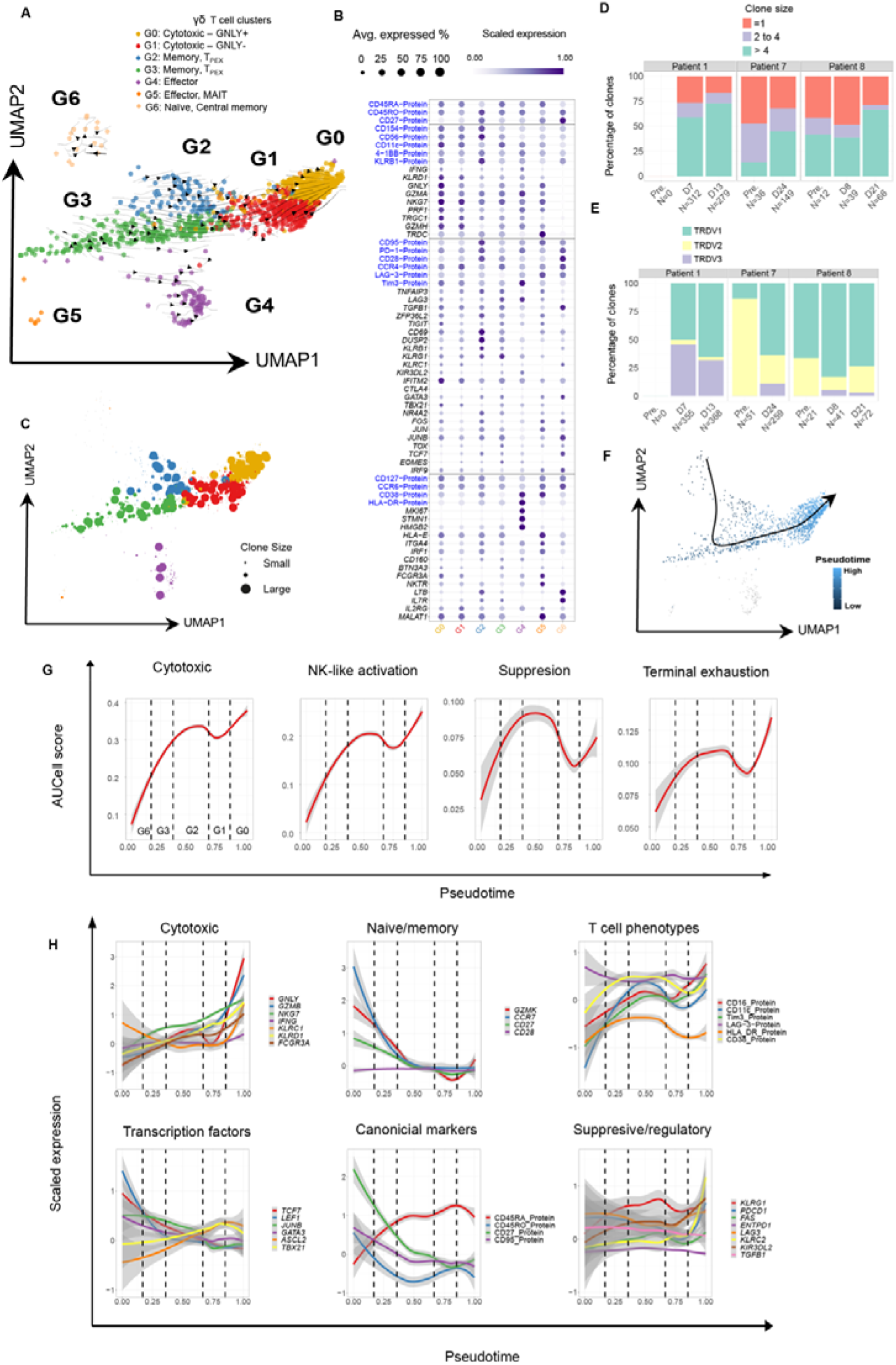
Clonally expanded γδ T cells revealed phenotypic plasticity and repertoire diversity. **A)** UMAP showing seven clusters and extrapolated future states (arrows) estimated by RNA velocity, applied to γδ T cells. **(B)** Dot plot of expression values for selected genes and proteins (AbSeq). Shown are gene markers with significant p-value <0.1 (using MAST, see methods). The size of each point represents the percentage of cells with non-zero expression. The colour represents the scaled log expression averaged over all samples. **(C)** Clone size mapped onto the UMAP plot, revealing their location across clusters. **(D)** Distribution of clone size, for each sample time point and patient. The number of clones are indicated by N. **(E)** Distribution of clones based on the gene usage of the delta variable chain, for each sample time point and patient (number of clones indicated by N). **(F)** Inferred differentiation trajectory using pseudotime across clusters G6, G2, G3, G1, and G0 as terminal state. **(G)** Loess curves of gene modules calculated by AUCell, vs scaled pseudotime values along the inferred trajectory. **(H)** Loess curve of scaled gene expression vs scaled pseudotime values along the inferred trajectory.

In contrast to the single cytotoxic cluster (C1) identified in CD8^+^ T cells, we identified two distinct clusters of cytotoxic γδ T cells, G0 and G1, with cluster G0 presenting higher expression of *GNLY, IFNG* and *GZMA* when compared to G1, despite similar protein expression profile consistent with a T_EMRA_ phenotype (CD45RA^+^, CD27^-^) and high expression of CCR4 (Fig. 4B). Clusters G2 and G3 comprised of effector memory (CD127^+^, CD45RO^+^), expressing *CD69*, *TCF7*, *FOS* and *JUN*, as well as exhaustion markers (*TIGIT*, LAG3, Tim3), suggesting evidence of T_PEX_. Cells in G0-G3 expressed CD56, CD154 (CD40L), KLRB1, and *TNFAIP3* (encoding A20), consistent with an NK-like signature^49, 50^. Similar to the C4 cluster in CD8^+^ T cells, γδT in G4 had a proliferating (*MKI67, STMN1, HMGB2*), activation (CD38, HLA-DR) and inhibitory (PD-1, TIM3) phenotype. Cluster G5 comprised of a small subset of cells resembling the effector MAIT phenotype observed in CD8^+^ T cells. Cluster G6 comprised a small subset of cells with a memory-like signature (*IL7R*). Most clusters expressed the butyrophilin gene *BTN3A3*, and HLA genes (Supplementary Table S5), which indicate interactions between γδ T cells via TCR mediated recognition of minor HLA molecules^17, 19^. In summary, γδ T cells comprised of similar phenotypes with the CD8^+^ T cells, including a cytotoxic NK-like phenotype shared between clusters G0-G3.

To investigate the intra-clonal phenotypic heterogeneity and cell fate of γδ T cells, full-length TCR sequences were used to identify clonally expanded subsets, which were then mapped onto the identified clusters. Full-length sequences were recovered for up to 80% of cells (Extended Data Fig. 7B). Clonotypes were observed in all samples and across all clusters (Fig.4C, Extended Data Fig. 7C), and with the clone size increasing from pre- to post-infusion (Fig. 4D). The majority of expanded clones identified at pre-infusion samples carried a TRDV2-TRGV9 genotype (Extended Data Fig. 7D), which are commonly found in circulation in humans^19, 50^, while clones at post-infusion were dominated by TRDV1 or TRDV3 genes (Fig. 4E), with TRDV1 paired with TRGV9, TRGV8, or TRGV4 (Extended Data Fig. 7D). TRDV1, TRDV2 and TRDV3 genotypes occupied all clusters (Extended Data Fig. 7E). Similar to CD8^+^ cells, we also found intra-clonal phenotypic heterogeneity within clones, revealing a transcriptional transition between clusters across time points (Extended Data Fig. 7F).

To study the dynamics between γδ T cell states, we performed similar trajectory analysis to the CD8^+^ T cells with RNA velocity (Fig. 4A), as well as by inferring cluster connectivity and differentiation trajectories using PAGA and CellRank (Extended Data Fig. 7, G and H). γδ T cells differentiated along a trajectory largely overlapping with the CD8^+^ T cells, originating from the naïve central memory subset (G6) and evolving through the memory-T_PEX_ clusters (G2/G3), and finally terminating towards the cytotoxic states (G1, G0) (Fig. 4F). The directionality of this trajectory correlated with the increased clonal expansion measured in cytotoxic clusters G0 and G1 (Fig. 4C). Transition probabilities between clusters confirmed the self-cycling nature of the highly proliferating cells in G4, with evidence of transition towards memory G3 and cytotoxic clusters G0 (Extended Data Fig. 7H).

Gene signature scores along the inferred trajectory revealed an increase in exhaustion, cytotoxic and NK-like profiles towards the terminal cytotoxic cluster (G0), while NK-like suppression peaked in the intermediate cell states (G2, G3) (Fig. 4G). Along this trajectory, transcription factors involved in memory differentiation such as *TCF7*, *GATA3*, *JUNB*, *NR4A2* declined, while *TBX21* increased (Fig. 4H).

Overall, these results revealed significant clonal expansion of unconventional γδ T cells predominantly due to an expanding TRDV1 clonotype, recapitulated known gene heterogeneity among γδ T cells^50^, and demonstrated a shared differentiation trajectory and phenotypic plasticity with CAR^+^ and CAR^-^ CD8^+^ T cells, which differentiated into a cytotoxic, NK-like and exhausted state.

### CAR^+^ and CAR^-^ T-cells are associated with altered serum cytokines levels following infusion

We next investigated whether the heterogenous clusters we discovered were associated with changes in serum cytokine levels, which were measured for the eight patients before CAR T cell infusion and at time-points within the first 30 days post-infusion^21, 22^. Comparison with pre-infusion baseline levels revealed an increase at post-infusion in pro-inflammatory cytokines IL-18, IL-12p40, and a decrease in MCP1 (encoded in *CCL2*) and homeostatic cytokine IL-15 (p-value<0.05, Fig. 5A). For the four patients with single cell multi-omics data (P1, P2, P7, P8), we assigned a gene-set score quantifying the degree to which each immune cell subset was expressed in pre- and post-infusion, including the CD8^+^ clusters, and calculated the Spearman correlation between these gene-set scores and measured cytokine levels to form an association score. This association score thus quantifies the degree to which cytokines simultaneously increases or decreases with each cluster gene signature. We also calculated this association between measured CAR^+^ T counts and clusters. Significant associations were found between pro-inflammatory cytokines IL-18, IL12p40, TNF-α, and IFN-γ, with γδT and NK cells (Fig. 5B), and with CAR^+^ T counts (Fig. 5B). These cytokines were also associated with CAR^+^ CD8^+^ T cells in clusters C1 (cytotoxic), C2 and C3 (memory T_PEX_) (Fig. 5C), but not with the corresponding CAR^-^ CD8^+^ subsets (Fig. 5D). These associations are consistent with the increased proportion of NK and NK-like CD8^+^ subsets in circulation, which are known to respond to innate-like cytokine stimulation such as IL-12 and IL-18 to make IFN-γ ^23–25^. In contrast, decline in homeostatic cytokine IL-15 was associated with a corresponding reduction of CAR^+^ and CAR^-^ CD8^+^ T cells in clusters C4 and C6 (Fig. 5, C and D), as well as monocytes and CD4^+^ cells (Fig. 5B).

**Fig. 5.**
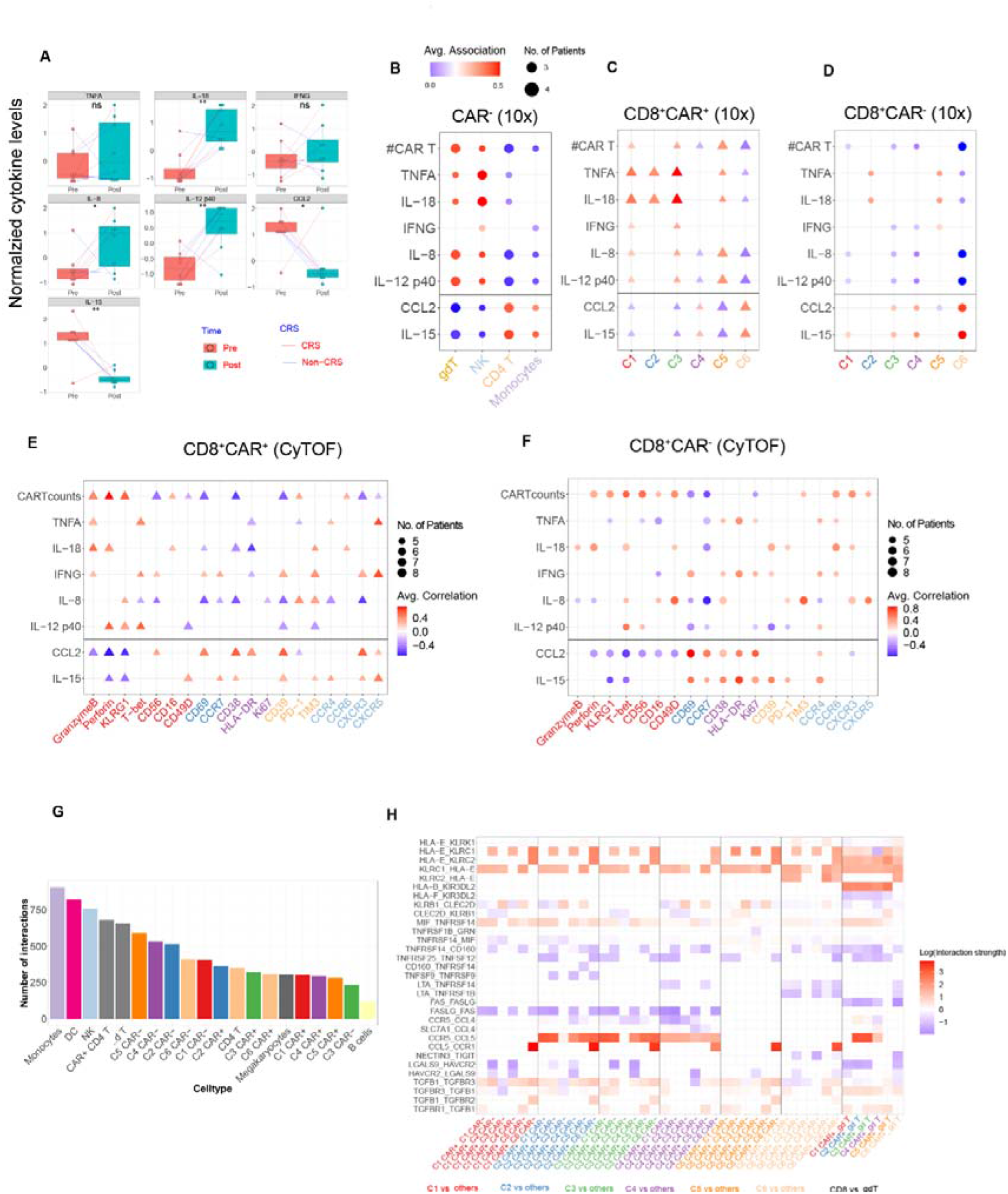
Serum cytokine levels are associated with subsets of patients’ and CAR^+^ T cells following infusion. **(A)** Box plot showing significant changes in cytokine levels from pre- to post-infusion (Wilcoxon test, p-value < 0.05 *, 0.01<p-value<0.05 **, <0.01 ***. Red lines represent 3 patients with low grade CRS, blue lines represent the remaining five patients. **(B)** Associations between changes in cytokine levels from pre- to post-infusion and gene signature scores characterising immune cell subsets. **(C-D)** Similar to (B) association between changes in cytokine levels from pre- to post-infusion with gene signature scores identifying clusters of CD8^+^ T cells from single cell multi-omics data in 4 patients. Shown are both CAR^+^ (C1-C6) (C) and CAR^-^ (C1*-C6*) clusters (D). Size of the symbols represent the number of patients with a significant association, colour represents magnitude of association, see Methods. **(E-F)** Association levels between changes in cytokine levels from pre- to post-infusion and mass cytometry protein expression levels in both CAR^+^ (E) and CAR^-^ (F) CD8^+^ T cells for all 8 patients. **(G)** Distribution of cell-cell interactions identified between CAR^+^ T cells and the patients’ immune cells utilizing only post-infusion samples. **(H)** Heatmap showing the cell-cell interactions between clusters of CD8^+^ CAR^+^ T cells with CD8^+^ CAR^-^ T cells and γδ T cells. Only cells from post-infusion samples are analysed. Each column represents interactions between two clusters.

We used mass cytometry to validate the cytokine-CD8^+^ T cluster associations on the remaining patients for CAR^+^ (Fig. 5E) and CAR^-^ CD8^+^ T cells (Fig. 5F). Pro-inflammatory cytokines (TNF-α, IL-18, IL12p40, and IFN-γ) were positively associated with CAR^+^ T cells with cytotoxic and effector markers (T-bet, perforin, and granzyme B), but also CD49D, CD56, and CCR6 which confirmed the associations with the NK-like signature from multi-omics data^23, 24^. In both the CAR^-^ and CAR^+^ T compartment, homeostatic cytokine IL-15 was associated with proliferation (HLA-DR, CD38) and exhaustion markers (CD39 (encoded in *ENTPD1*), PD-1, and Tim-3), (Fig.5, E and F). Finally, CAR^+^CD8^+^ T cells expressing chemokine receptors (CXCR3, CXCR5) were positively associated with IL-15, which were found expressed on the memory -T_PEX_ and proliferative T cells subsets (Fig. 5E).

We next investigated whether the association between the cell populations and the cytokine levels were consistent with cell-to-cell interactions between endogenous CAR^-^ immune cells and CAR^+^ T cells, for instance through cytokine-cognate receptor interactions. To test this hypothesis, we predicted the cell-to-cell interactions between the circulating immune cells and CAR^+^ T cells by computing statistically significant interactions mediated by chemokines, cytokines and HLA molecules following infusion using CellphoneDB^51^. We identified a total of 9143 interactions (Fig. 5G) between CAR^+^ T cells and other immune cells. These cells interacted predominantly with CAR^+^ T cells in C4, C5, and C6 via HLA-E expression (Fig. 5H). CAR^+^ CD8^+^ T cells in the clusters of memory T_PEX_ (C2) and proliferative subset (C4) were predicted to interact with endogenous CD8^+^ and γ T cells via chemokine receptors CCR5 (Fig. 5H). A regulatory set of interactions was also discovered between CAR^+^ and CAR^-^ T cells, via expression of co-inhibitory molecules such as KLRB1-CLEC2D, TIGIT-NECTIN3, HAVCR2 (Tim-3)-LGALS9, TGFB, and TNF receptors such as TNFRSF25-TNFRSF12 (Tweak), and TNFRSF14, which interact with multiple ligands influencing both activation and regulation^52^ (Fig. 5H).

Together, these data revealed associations between NK/NKT and NK-like CD8^+^ T cells with pro-inflammatory cytokines, including the innate-like IL-18 and IL-12, while proliferating and activated CD8^+^ T cells were associated with homeostatic cytokine IL-15. Lastly, cell-cell interactions with CAR T cells contribute to homing and regulatory functions of recovering immune cells following infusion.

## DISCUSSION

This comprehensive multi-omics analysis performed on longitudinal samples identified unexpected shared molecular and phenotypic features between CAR^+^ and CAR^-^ CD8^+^ T cells, which were used to characterize the differentiation process in the early phase following infusion. We found that CAR^+^ T cells respond to additive signals, which include CAR-specific stimulation due to aberrant CD19^+^ B cells^21^ as well as stimuli that are shared with other T cells, such as homeostatic proliferation induced by lympho-depletion, TCR-dependent stimulation and cytokines, e.g., the innate-like interleukins IL-12 and IL-18 (Graphical Abstract). This NK-like profile is associated with the rapid increase in circulating pro-inflammatory cytokines. These stimuli induce a shared differentiation trajectory that describe both infused CAR^+^ T cells and patients’ T cells. The combination of additive stimuli leads to a rapid differentiation into a poly-functional cytotoxic response, which includes an innate NK-like subset within both CAR^+^ and CAR^-^ CD8^+^ T cells. The endogenous repertoire of CD8^+^ T cells undergo greater clonal expansion, likely due to pre-existing memory clones that survived lympho-depletion and antigen stimulation. In contrast, CAR^+^ T cells develop higher proliferation, exhaustion, and metabolic activities in line with the high antigen stimulation due to the addition of CAR as previously reported^11, 13, 53^.

We identified a terminal state of differentiation with a polyfunctional cytotoxic profile which included an NK-like signature that was common across γδ, and CD8^+^ T cells. Strikingly, this cell state was shared across both CAR^+^ and CAR^-^ CD8^+^ T cells, expressed CD56, and was associated with clinically relevant phenotypes, specifically increasing levels of cytokines IL-12 and IL-18. NK-like CD8^+^ T cells were also recently reported in CAR^+^ T cells under persisting stimulation *in vitro*^15^. The NK-like profile identified in this *in vivo* study had a low expression of transcription factors *SOX4,* and *ID3,* which were identified as the main signature of the NK-like subset identified *in vitro*^15^. Instead, NK-like cells detected in our study expressed transcription factors *KLF2*, *KLF3*, and *MYBL1*, which were detected in cytotoxic, non-dysfunctional subsets in tumor infiltrating T cells^29^, as well as in CAR^+^ T cells *in vitro*^15^. A previous study using scRNA-seq data generated from autologous CAR^+^ T cells circulating at about a month after infusion, revealed a proliferative and exhausted profile, but no clear evidence of an NK-like signature^13^. In contrast to previous studies, our single cell multi-omics approach was designed to address the differentiation process of CAR^+^ T cells and how the cell states differ from the patients’ immune cells, thus providing an accurate identification of the phenotypic heterogeneity of patients’ and CAR^+^ T cells in early phase of therapy.

This study provided a molecular and phenotypic description of the differentiation process and cell fate dynamics within the γδ T cells. We reported large clonal expansion across a diverse TCR repertoire, which revealed an unexpected increase in non δ2 subsets, which is the most common subset found in circulation. Contrary to the well-studied γ9δ2 subset, the δ1 and δ3 repertoires are diverse in the antigenic spectrum and are known to be involved in cytotoxic responses against pathogens^17, 18, 54^, as well as in regulatory functions^55^. We observed an imbalance of δ1 and δ2 in circulating γδ T cells, which has been previously shown to be associated with tumor invasiveness^55^, and have been reported in the peripheral blood of patients with chronic viral infections and patients with leukemia^56, 57^. Our results provide relevant discoveries for future studies investigating the use of γδ T cells for cellular therapies58, such as the molecular patterns associated with the evolution of γδ T cells following lympho-depletion and the fate that these cells acquire and share with conventional T cells and CAR+ T cells.

The results of this work are also of relevance for translational applications, for instance to better understand the outcomes of donor derived CAR^+^ T cells in the patient following infusion. Donor-derived products are considered viable solutions to extend CAR^+^ T cell treatments to a broader range of patients, with the additional advantage of simplifying or accelerating manufacturing of products^59, 60^. In this unique trial, we had the opportunity to study donor derived CAR T cells in patients that shared the same genetic background of endogenous cells (engrafted following HSCT from the same donor), and provided evidence that despite the CAR manufacturing, the evolution of genetically similar patients’ immune- and CAR^+^ T-cells followed a similar path and shared similar phenotypes. We did not observe significant differences in the patient (A1) treated with autologous CAR, which again confirmed that the matched donor conditions prevented allogenic responses against CAR^+^ T cells. Furthermore, we did not observe significant differences in T cell phenotypes in patient P3, who had a rejection of the graft at the time of therapy. These findings suggest that the differentiation process undertaken by T cells is not affected by the genetic differences between the donor and the recipient prior to infusion, although extended studies on allogenic CAR T cells are needed.

Our study also provided insight into the proliferation of CAR^+^ and immune cells following immune-depletion. During the early phase of lymphopenia following immune-depletion, infections are common and can affect persistence of both CAR^+^ T ^5^ and patient’s immune cells, for instance by limiting T cell diversity or driving severe inflammatory responses^61^. Immune-depletion has also been shown to drive homeostatic proliferation in CD19 CAR^+^ T cells^62^. For CAR-CD8^+^ T and γδ T immune cells, we found significant clonal expansion from memory cells present in the patient’s blood prior to infusion. These findings are consistent with the increased proliferation induced by lympho-depletion, and are also supported by transcriptomic profiles revealing activation of TCR signalling pathways. Several of these clones were found to match viral specific TCR sequence, and notably similar findings were found for CAR^+^ T cells, that underwent clonal expansion from a diverse repertoire in the IP, notably a significant expansion of a CMV-specific CAR^+^ T cells in P7 dominating the circulating population. However, CAR^+^ T cells were found less clonally expanded compared to non-CAR T cells, therefore other mechanisms are at play in controlling clonal expansion. Indeed, many large memory clones were found pre-infusion, thus suggesting that preferential expansion may have occurred due to competition for niches such as cytokines and bystander activation^63^. It is known that memory responses to viral infections, such as CMV and EBV and influenza, affect immune reconstitution in the context of both HSCT and cellular therapy^64, 65^, and these dynamics could also influence CAR^+^ T cell proliferation and expansion. Further studies could monitor CAR^+^ T cells in larger cohorts and for longer periods of time to further investigate these mechanisms.

The findings of this work provide researchers with a better understanding of CAR^+^ T cell differentiation, as well as potential solutions to utilise this knowledge and improve CAR^+^ T cell therapies. For instance, we identified associations of a terminally differentiated state of NK-like CD8^+^ subset with pro-inflammatory cytokines. This phenotype is known to contribute to inflammatory response, by secretion of IFN-γ via TCR independent stimuli^23, 24, 49^. CAR^+^ T cell products which will result in a reduced inflammatory response, for instance, can thus be obtained by enriching for less differentiated subsets of T cells, reduce affinity of CAR for antigen targets, or the use of pharmacological therapies to slow down cell proliferation and activation.

One limitation of this study is the small sample size in this trial, and our findings should be validated, in larger cohorts, for instance the association between cytokines and subsets of CAR^-^ and CAR^+^ T cells. Single cell multi-omics was used to identify molecular and cellular subsets which can now be directly measured with easier and more affordable technologies, hence confirming their predictive power.

In this cohort, two patients have developed CAR^+^ T cell induced lymphomas^22^. The analysis of multi-omics data did not reveal any differences between the two patients (P2 and P8) compared to the others. These two lymphomas were diagnosed at a later time point in both patients, and well beyond the first month post-infusion^22^. The CAR^+^ T cell induced lymphoma in both P2 and P8 were monoclonal in the endogenous TCR, with the P2 clone being a CD4^+^ CAR^+^ T cell which we successfully identified in a minority of cells at day 24 post-infusion (not shown), and similarly for P8 only one cell in CAR^+^ T cell sample at day 8 carried the same TCR as the lymphoma. Further investigations are underway to elucidate the mechanisms that underlie these conditions.

This comprehensive multi-omics data set enables future work to identify additional cell and gene signatures to accelerate mechanistic and functional investigations into the role of specific genes and host immune cells in CAR T cell therapies. New biomarkers are needed to identify patients that are likely to relapse and understand what determines CAR^+^ T cell expansion and persistence.

## MATERIALS AND METHODS

### Study design

The purpose of this study was to understand the evolution of CAR^+^ T cells and CAR^-^ immune populations circulating in patients during the initial stage of therapy. To this end, we generated and analyzed 20 multi-omic datasets comprised of 11 PBMC populations, 8 CAR^+^ T cell populations and one healthy PBMC reference population (publicly released by 10X Genomics) (Supplementary Table S2). We also compiled mass cytometry (CyTOF) data which comprised of 36 PBMC samples and 8 infusion products.

### Subjects and samples

All samples were collected from Westmead Hospital, with written informed consent obtained from all participants in the CARTELL study (ACTRN12617001579381). The relevant ethical approvals related to the CARTELL study are from the NHMRC (4089 AU RED HREC/14/WMEAD/332) and the University of New South Wales (reference number HC190012). Samples consisted of cryopreserved PBMC isolated by density gradient centrifugation.

### Cell preparation for scRNA-seq

Cryopreserved cells were thawed and transferred to room temperature RPMI 1640 media (Gibco). Subsequent washes were performed with PBS containing 1% BSA. Manual cell counts were performed with trypan blue and C-Chip haemocytometer slides (NanoEntek). Samples were divided into aliquots containing 1 million cells and designated for either CAR19^+^ or PBMC population analysis. Cells were pelleted by centrifugation and incubated with human Fc Block (BD Pharmingen), except for P1 day 13, P2 day 24, and P2 product. Cells were next stained with a cocktail of oligonucleotide-conjugated antibodies (Supplementary Table S1) for 45 minutes at 4°C. For samples to be enriched for CAR19^+^ T cells, additional staining with fluorophore-conjugated antibodies for CD3, CAR19, CD19, CCR7, CD8, PD-1, and CD45RA was performed after 25 minutes of initial incubation with oligonucleotide-conjugated antibodies followed by three washes following incubation. Samples for CAR19^+^ analysis underwent additional viability staining (Fixable Yellow/7-AAD/propidium iodide [Invitrogen]) according to manufacturer’s instructions prior to sorting. Samples for PBMC analysis proceeded directly to single-cell library preparation. Sorting for the CAR^+^ population (lymphocytes/singlets/live cells/CD19^-^/CD3^+^/CAR19^+^ cells) was performed on a BD FACSAria III (BD Biosciences) at the UNSW Flow Cytometry core facility with single cell precision into 15 mL centrifuge tubes. Compensation was performed with single stained compensation beads and calculated using the associated FACSDiva software (BD Biosciences). Following sorting, cells were pelleted, and a manual cell count was performed.

### Single-cell library preparation and sequencing

Single-cell library preparation steps using the 10X Genomics platform were performed using Chromium Single Cell 3’ Reagents Kits v2 (10X Genomics, PN-120267) and Chromium Single Cell 3’ Reagents Kits v3 (10X Genomics, PN-1000092). An additional AbSeq PCR1 primer (2 μl) (BD Biosciences, cat. No. 91-1086) was added to the cDNA amplification master mix to enable amplification of the AbSeq oligonucleotide-conjugated antibodies. Following cDNA amplification, single-cell gene expression libraries were purified using SPRIselect beads (0.6X) (Beckman Coulter). The supernatant (containing the AbSeq libraries) was retained, and an additional SPRIselect bead incubation was performed (1.8X). AbSeq libraries were eluted in 25 μlof buffer EB. AbSeq sample index PCR was performed using 11 μl of AbSeq products, 2 μl SI-PCR primer (from 10X 3’ v2 kit), 2 μl reverse indexing primer (supplied from BD Biosciences), 25 μl 2X PCR mastermix (supplied from BD Biosciences), and 10 μl water. Reactions were incubated: denaturation 98°C for 45s, PCR cycling of 98°C for 20s, 54°C for 30s, 72°C for 20s, and final extension 72°C for 1 minute. Between 10 to 15 cycles were used based on input cell numbers. Following sample index PCR, AbSeq libraries were purified with a double-sided SPRIselect bead size selection (0.6X and 1.0X).

Single-cell library preparation steps using the BD Rhapsody platform were performed according to the manufacturer’s protocol for ‘mRNA Whole Transcriptome Analysis (WTA), AbSeq, and Sample Tag Library Preparation’ (Revision 23-21752-00). Single-cell library preparation for CAR T cell products from patients P1, P7, and P8 were performed using the BD Rhapsody platform. All other scRNA-seq libraries were generated on the 10X single-cell platform.

Sequencing libraries were quality controlled using a combination of agarose gel electrophoresis, Quant-iT™ PicoGreen™ dsDNA Assay (Invitrogen) quantification and running on a LabChip GX platform (Perkin Elmer) using a High Sensitivity Assay (CLS760672). Gene expression and AbSeq libraries were pooled and sequenced on a combination of Illumina NextSeq500 and NovaSeq6000 platforms at the UNSW Ramaciotti Centre for Genomics using recommended read lengths.

### Immune receptor sequencing RAGE-seq

Full length TCR and BCR were obtained from 10X 3’ library via the RAGEseq methods, as previously described^45^. Briefly, between 50-500ng of full-length cDNA generated from the Chromium Single Cell 3’ protocol (10X Genomics) was used for targeted enrichment of all functional TCR and BCR genes following the Roche-NimbleGen double capture protocol. Briefly, cDNA libraries were incubated overnight at 47 °C with probes and hybridization reagents (SeqCap EZ Accessory Kit v2; SeqCap EZ Hybridisation and Wash Kit) and then washed and hybridized a second time overnight for further enrichment. Following each round of hybridisation and capture, PCR was performed using KAPA HotStart HIFI with 1 µM 10X_F primer (AAGCAGTGGTATCAACGCAGAGT) and 1µM 10X_R primer (CTACACGACGCTCTTCCGATCT) with the following conditions: 98 °C for 3 min; [98 °C for 20 s, 65 °C for 15 s, 72 °C for 1 min 30 s] × 5 cycles (first round) or × 20cycles (second round); 72 °C for 3 min. Post-capture cDNA library size ranged from 0.6 to 2 kb.

Post-capture cDNA libraries were prepared for long-read sequencing using Oxford Nanopore Technologies’ (ONT) 1D adapter ligation sequencing kit (SQK-LSK109 or SQK-LSK110) and sequenced on R9.4.1 PromethION flow cells (FLO-PRO002) were base-called using Guppy (v3 or v4). The resulting base-called FASTQ files were de-multiplexed by 10X cell barcodes, using a direct sequence matching strategy described previously^45^. Briefly, forward and reverse-complemented cell-barcode sequences (16 nt) were used to demultiplex the nanopore sequencing reads by scanning the first and last 200 nt of any read longer than 250 nt for a matching sequence, with < 2 mismatches. All reads assigned to a given cell were then de novo assembled using Canu (v1.8) and sequence-polished using Racon (v1.3.3), then polished contigs were analysed with IGBlast, as described previously.

### Pre-processing of single-cell RNA-seq and AbSeq data

Gene expression matrices corresponding to UMI counts from Rhapsody scRNA-seq libraries were produced using the BD Seven Bridge Genomics platform. Matrices from 10X scRNA-seq libraries were produced using the Cell Ranger workflow (v3.0.1, 10X Genomics). We utilised a modified GRCh38 reference genome which was appended with a sequence that corresponded to the transcript product of the CAR T transgene. For the 10X samples, a custom script obtained from the AbSeq manufacturer (BD® bioinformatics) was applied to the protein expression base call files to obtain the protein expression matrix. The healthy control sample was downloaded from the 10x website (https://www.10xgenomics.com/resources/datasets).

Samples were quality controlled to remove cells with low mitochondrial content, and low/high number of unique genes and total counts. Gene expression was normalized using scran with default parameters, while protein expression normalized using centred-log-ratio (CLR) normalization. Potential doublets were removed using the function doubleCluster in the R package scran^12^.

### Mass cytometry (CyTOF) data

Paired cryopreserved CAR19 T cell product pilot vials and donor PBMCs were phenotyped by mass cytometry using metal-conjugated mAbs. All antibodies were validated, pre-tittered and supplied in per-test amounts by the Ramaciotti Facility for Human Systems Biology Mass Cytometry Reagent Bank, Sydney, Australia. Metal-conjugated mAb were purchased from Fluidigm. Unconjugated mAbs were purchased carrier-protein-free and conjugated by the Ramaciotti Facility for Human Systems Biology with metal isotopes using the MaxPar X8 labelling reagent kits (Fluidigm), according to the manufacturer’s protocol. Viability staining, barcoding of paired samples with unique mass labels, staining with surface antigen-specific mAbs, fixed and stained with intracellular antigen-specific mAbs before data acquisition with a CyTOF 2 Helios upgraded mass cytometer (Fluidigm) was performed. Data collected in FCS3 file format was normalised for signal intensity of EQ Four Element Calibration Beads using the Helios software (V6.3.119). FlowJo 10.4.2 software (FlowJo, LLC) was used to identify CAR19 T cells and donor T cells as comparison, based on the signal of the CD45 barcoding antibodies.

### Clustering and identification of T cell subsets

Dimensionality reduction and clustering for the scRNA-seq dataset was performed using the Seurat R package^66^. Gene expression matrices was aggregated using the FindIntegrationAnchors and IntegrateData functions. The top 3000 highly variable genes were extracted using the FindVariableFeatures function. Principal component analysis was run using the RunPCA function with the option npcs=30 following by neighbour and cluster calculation using the FindNeighbours and FindClusters functions respectively. We explored resolution parameters in the range of 0.01 to 0.6 to determine the marker genes and proteins which adequately explained the observed clusters.

We used these protein and gene markers to classify canonical immune cell types: CD8^+^ T cells (CD8^+^, CD3^+^, CD4^-^, *CD8A*^+^), γδT cells (CD3^+^,CD4^-^, CD8^-^, *TRDC*^+^, *TRGC1*^+^), natural killer (NK) and NKT cells (CD8^-^, CD56^high^, CD16^+^, *NKG7*^+^), CD4^+^ T cells (CD4^+^, CD3^+^, CD8^-^), monocytes (CD14^+^, CD3^-^, *CD14*^+^), dendritic cells (DC) (CD3^-^, CD14^-^, CD16^-^, *HLA-DRB1*^+^), B cells (CD3^-^, *JCHAIN*^+^, *ITM2C*^+^) and megakaryocytes (*NRGN*^+^).

### Differential expression analysis

Two differential expression analyses were performed depending on whether the analysis was performed within the same sample to find heterogenous clusters, or between different samples to account for inter-sample differences. In the first approach, for each sample, DGE and DPE (intra-sample) analysis were both performed using the FindAllMarkers function, with the “MAST” ^67^ and “Wilcoxon sum rank test” test options respectively. The clusters used in this analysis were formed from the integrated data as previously described. Differentially expressed genes and proteins that appeared in the majority of samples were designated marker gene and proteins. This allowed identification of markers which truly represented the clusters across most samples, in contrast to markers which were only differentially expressed in one sample. UMAP was then used to visualize the clustering results, using nearest neighbour=30 and min_dist=0.01. The dotplots were generated using ggplot2, with the averages obtained over all samples across all patients. In the second approach, DGE and DPE (inter-sample) analysis were performed using the edgeR wrapper run_de function in the Libra R library^68^ with the likelihood-ratio (LRT) and pseudobulk option, with the patients serving as the replicates.

For the mass cytometry data, integration was performed using the FindIntegrationAnchors and IntegrateData, and performed separately for P1, P2, P7, P8 in one group, and P3-P6 for the second group. DPE analysis was performed using the FindAllMarkers function using the “Wilcoxon sum rank test” test. UMAP was used to visualize clustering, with nearest neighbour=30 and min_dist=0.1.

### Gene set analysis

Gene set enrichment analysis (GSEA) was performed using the fgsea function in the fgsea R package ^69^ with default parameters, based on DGE results ranked by the fold-change. Pathways and gene sets were downloaded from the MSigDB^70, 71^ in combination with curated T cell specific gene-sets (Supplementary Table S8). Average NES values were obtained by averaging over the NES values over each sample. AUcell ^72^ with default parameters was used to assign gene signature scores.

### Cell cycle analysis

Cell cycle scoring and annotation was performed using the CellCycleScoring function in the Seurat package.

### Trajectory analysis and RNA velocity

The PAGA plots were generated by running pca (scanpy:pca), finding the nearest neighbours (scanpy:neighbours) and running PAGA (scanpy:paga) using default parameters on the integrated gene expression matrix, and using the pre-defined CD8^+^ T and γδT clusters.

Slingshot was applied to the UMAP formed from the integrated gene expression matrices using the getLineages and getCurves functions in the slingshot package^73^, and by manually assigning an initial root value. The pseudotime values generated from Slingshot were used to generate the smoothed gene expression and AUcell vs scaled pseudotime curves, where the smoothed expressions were calculated using the *geom_smooth()* R function with default parameters and the pseudotime was scaled to take on values between 0 and 1. Further scaling was performed to ensure the endogenous and CAR T psuedotime values corresponded to the same clusters.

RNA velocity was implemented by first extracting the unspliced and spliced matrices generated from the 10x data using bustools ^74^, and then running the standard RNA velocity pipeline (scvelo: filter_and_normalize, moments, recover_dynamics, velocity) using default parameters, and the “dynamical” mode^48^. The final RNA velocity plot was generated by concatenating the individual RNA velocity plots.

The Cellrank python package^25^ was used to calculate the cluster transition probabilities using the transition_matrix function on each sample, with the result obtained by averaging across the samples.

### T Cell Receptor sequencing and analysis

T cell receptor α-β CDR3 amino-acid sequences were extracted using RageSeq^45^ on all samples except for the product samples. Analysis was performed with the bioinformatics pipeline proposed in the original manuscript with only minor modifications. Code available upon request. For the post-infusion PBMC samples, clones with at least one cell expressing the CAR T transgene were removed. This did not significantly affect any samples, except for the day 24 timepoint from Patient 3 which contained a larger proportion of CAR T cells than the other samples. T cell receptor γδ analysis followed the same procedure.

### Identification of known TCR specificities

To identify TCR sequences with known specificity, we downloaded all the available paired TCRαβ CDR3 sequences from the VDJdb data base^46^. We then used the Levenshtein distance to compute pairwise distances between TCRαβ CDR3 sequences and considered a close match only those with a pairwise distance ≤ 4.

### Association Analysis

The association between the CD8^+^ T clusters/canonical cell types with the cytokines was calculated by averaging across the correlation coefficients between cytokines and genes. Specifically denote X^k^ as the *N* x 2 matrix for the *k*th patient, where *N* is the number of genes which represents the cluster of interest, and the entries X^k^_i1_ and X^k^_i2_ denotes the average gene expression across all cells of the *i*^th^ gene at the pre- and post-infusion sample respectively. The genes were chosen which were differentially expressed (FC>1, p-value<0.1) in more than 9 samples. Denote also Y^k^ as the *M* x 2 matrix where *M* is the number of measured cytokines with the matrix entries Y^k^_i1_ and Y^k^_i2_ representing the measured cytokine levels at pre- and post-infusion respectively. We first calculate Z^k^ = corr(X^k^, Y^k^), which is the *M* x *N* Spearman correlation matrix between X^k^ and Y^k^. Then the final association between the *i*th cytokine and the cluster is given by (1/*NP*) 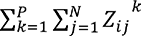 where *P*=4 is the number of patients used in the analysis. A similar procedure was used to calculate the association values within the CAR T cell subsets, using the product and circulating samples as time points. For the mass cytometry (CyTOF) data, associations were calculated for all the 8 patients, averaging the protein expressions across cells, and then utilising the same formula as per the genes.

### CellphoneDB

Cell-cell interactions based on the expression of known ligand-receptor pairs was calculated using CellPhoneDB version 2 using default parameters^51^. Only interactions within the top 5% quantile of the predicted mean values (mean value = 0.268), and a p-value < 0.01. This analysis is asymmetric, therefore we tested pairs of interactions twice.

## Data Availability

All data produced in the present study are available upon reasonable request to the authors

## DATA AVAILABILITY

All the single cell data of gene and protein expression are deposited with GEO XXXX. TCR sequences identified in this study are available upon request.

## Acknowledgments

This work was supported by the National Health and Medical Research Council, Australia (Project Grants 1102172, 1121704,1121643) and by a Translational Program Grant from Cancer Council NSW and the Cancer Institute NSW (TPG 19-01). FL was an NHMRC Career Development Fellow (1128416). EB was a Cancer Institute NSW Postgraduate Fellow and recipient of a NSW Ministry of Health Early/Mid-Career Cell and Gene Therapy Fellowship. HMM was previously an NHMRC postdoctoral fellow (GNT1037298) and is currently supported by the International Society for the Advancement of Cytometry (ISAC) Marylou Ingram Scholars program. We thank Katherine Jackson for helping with the bioinformatics analysis and for providing useful suggestions.

## Author contribution

FL conceptualise the study, led the data analysis and interpretation, and funded the study. RL conducted the bioinformatics analysis. CC performed the single cell multi omics experiments with help from TA. FL and RL wrote the manuscript, with contribution from CC. ID, JF, KJ, and TGA contributed to the TCR analysis. MS performed experiments for RAGE-seq. HM performed CYTOF experiments. KM led the clinical trial study and provided samples. DG, DB, KG, HMM, and EB contributed reagents, performed the experimental work and helped with data interpretation. All the authors have read the manuscript.

---

**Extended Data Fig. 1.**
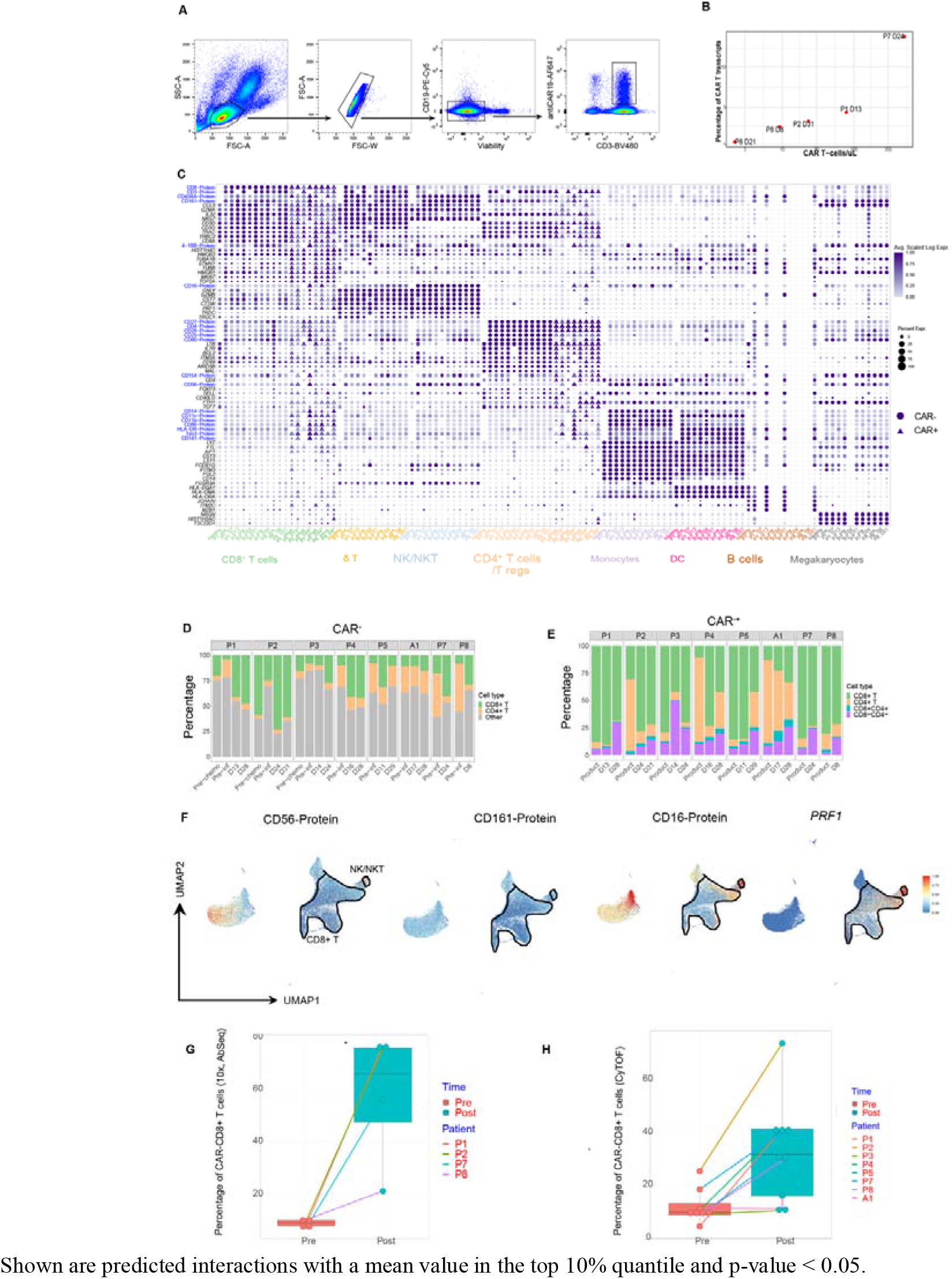
Single cell gene and protein expressions reveal endogenous (PBMC) and CAR T immune cell subsets. **(A)** Gating strategy to identify CAR^+^ T cells from total PBMC using flow cytometry cell sorting. (**B**) Scatter plot showing percentage of cells with detectable CAR T transcripts versus the number of CAR T cell counts in the blood via DNA quantification. (**C**) Dot plot of selected genes (scRNA-seq) and proteins (AbSeq) for each sample analysed via single cell multi-omics. The size of each point represents the percentage of cells with non-zero expression. The colour represents the scaled log expression (see Methods). CAR^-^ T cells are represented by filled circles, CAR^+^ T cells by filled triangles. (**D**) Stacked bar plot of the CD8*^+^* T and CD4*^+^* CAR^+^ T cells in each sample analysed by mass cytometry. (**E**) Stacked bar plot of the CD4*^+^* and CD8*^+^* subsets in each CAR^-^ sample analysed by mass cytometry. (**F**) UMAP with selected marker genes and proteins, identifying NK-like CD8+ and unconventional NK and NKT cells. (**G-H**) Proportion of CAR^+^CD8^+^ T cells identified with Abseq (N=4 patients. G) and Mass-cytometry data (N=8 patients, H).

**Extended Data Fig. 2.**
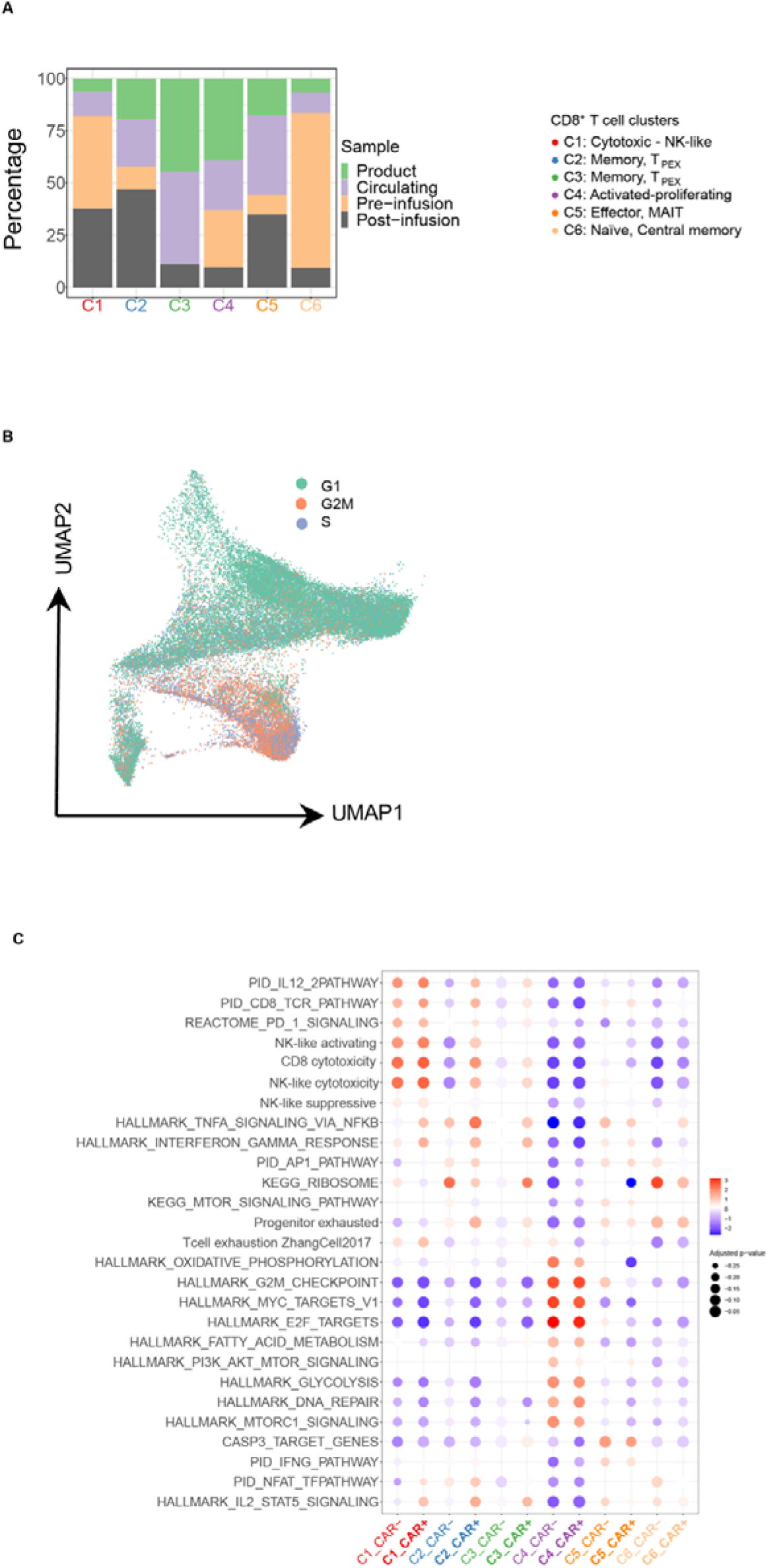
Characterization of CAR^+^ and CAR^-^ CD8^+^ T cells in clusters identified using single-cell multi-omics. **(A)** Distribution of sample type composition at each CD8^+^ cluster for both CAR^-^ and CAR^+^ T cells. **(B)** Projection of predicted G1, G2M and S phase cells on UMAP. **(C)** Dot plot of NES values from GSEA performed utilising cluster genes. Shown are both CAR^-^ and CAR^+^ T cells. Only significant gene sets are shown (p-value<0.05). The size of each point represents the –log10(p-value). The colour represents the NES. Gene sets were either taken from the MsigDB database or manually curated (Table S3).

**Extended Data Fig. 3.**
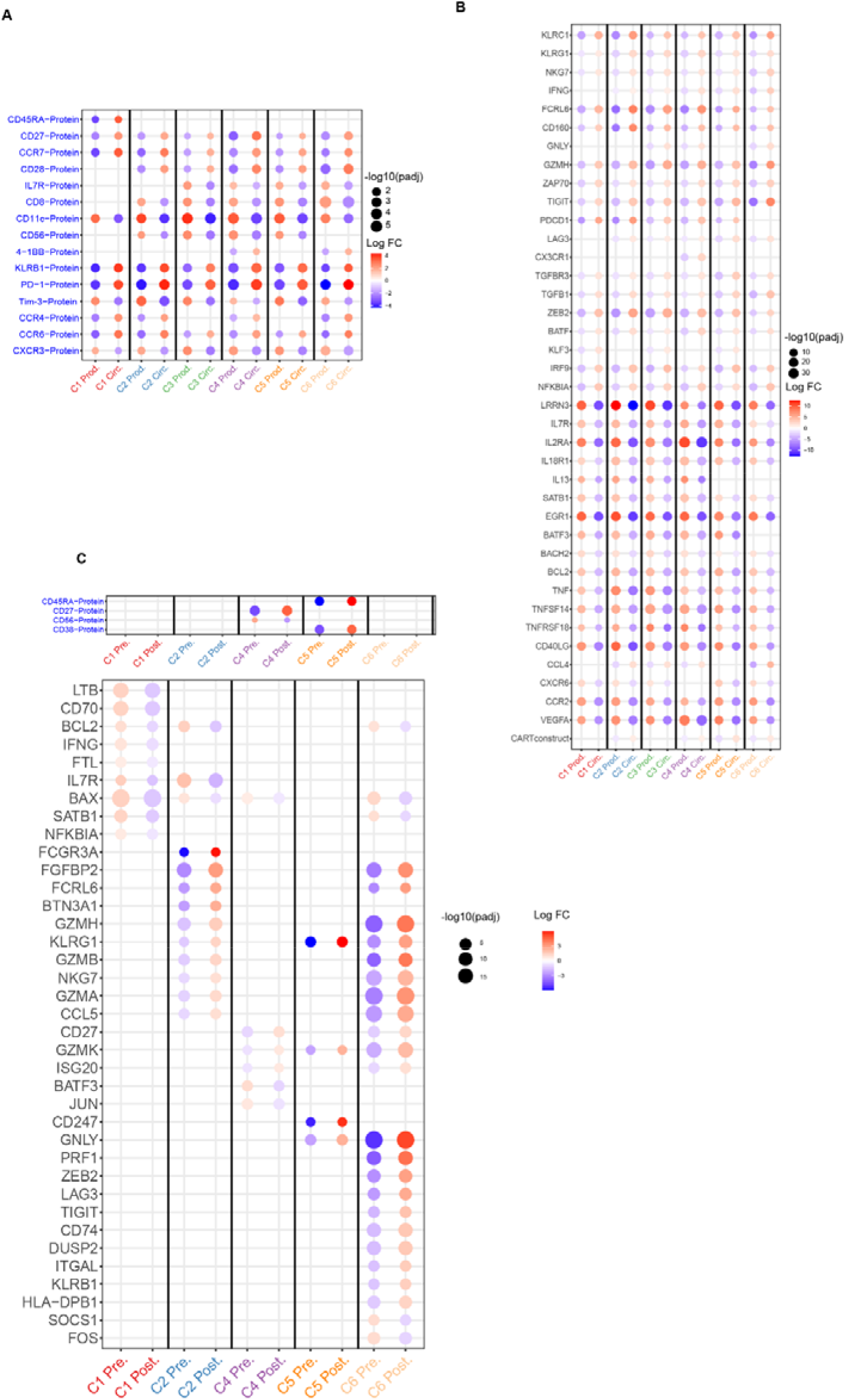
Differential gene and protein analysis between product vs circulating post-infusion and pre- vs post-infusion CD8^+^ T cells. **(A-B)** Dot plot of fold change values for gene (A) and protein (B) comparing product vs circulating CAR^+^ CD8^+^ T cells, calculated using edgeR with the patients as replicates. Shown are markers with significant p-value (<0.1 using edgeR). The size of each point represents –log10(adjusted p value). The colour represents the log fold change. **(C)** Dot plot of fold change values comparing pre vs post CAR^-^ CD8+ T cells, calculated using edgeR with the patients as replicates. Shown are markers with significant p-value (<0.1 using edgeR). The size of each point represents –log10(adjusted p value). The colour represents the log fold change.

**Extended Data Fig. 4.**
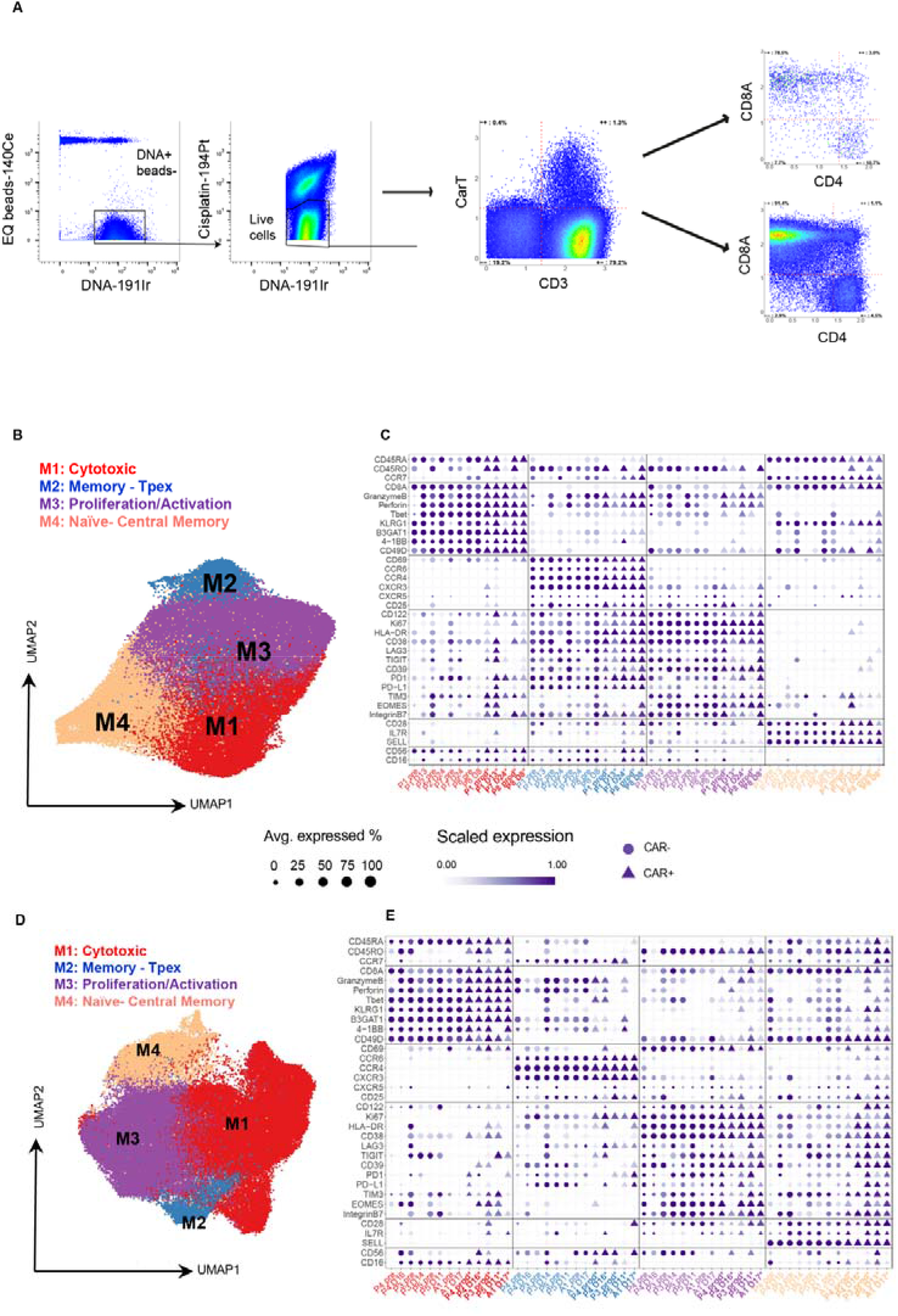
Characterization of CAR^+^ and CAR^-^ CD8^+^ T cells in clusters using mass cytometry. **(A)** Gating strategy to identify CAR^+^ T cells from total PBMC using mass cytometry. (**B**) UMAP of CD8*^+^* T cells and for both CAR^+^ and CAR^-^ T cells using mass cytometry, revealing four clusters. This plot corresponds to samples obtained from the same patients with available single cell multi-omics data, i.e., P1, P2, P7, P8. **(C)** Dot plot of scaled expression values (Materials and methods) for mass cytometry markers for the same patients as (B), and for both CAR^+^ and CAR^-^ T cells. Shown are markers with significant p-value (<0.1 using Wilcoxon Rank Sum test). The size of each point represents the percentage of cells with non-zero expression. The colour represents the scaled log expression. Endogenous samples are represented by filled circles, CAR T samples by filled triangles. **(D)** UMAP of CD8*^+^* T cells and for both CAR^+^ and CAR^-^ T cells, showing four clusters using data from patients P3, P4, P5, and A1. **(E)** Dot plot of scaled expression values (Materials and methods) for mass cytometry markers corresponding to the clusters identified in (D). Shown are markers with significant p-value (<0.1, Wilcoxon Rank Sum test). The size of each point represents the percentage of cells with non-zero expression. The colour represents the scaled log expression.’

**Extended Data Fig. 5.**
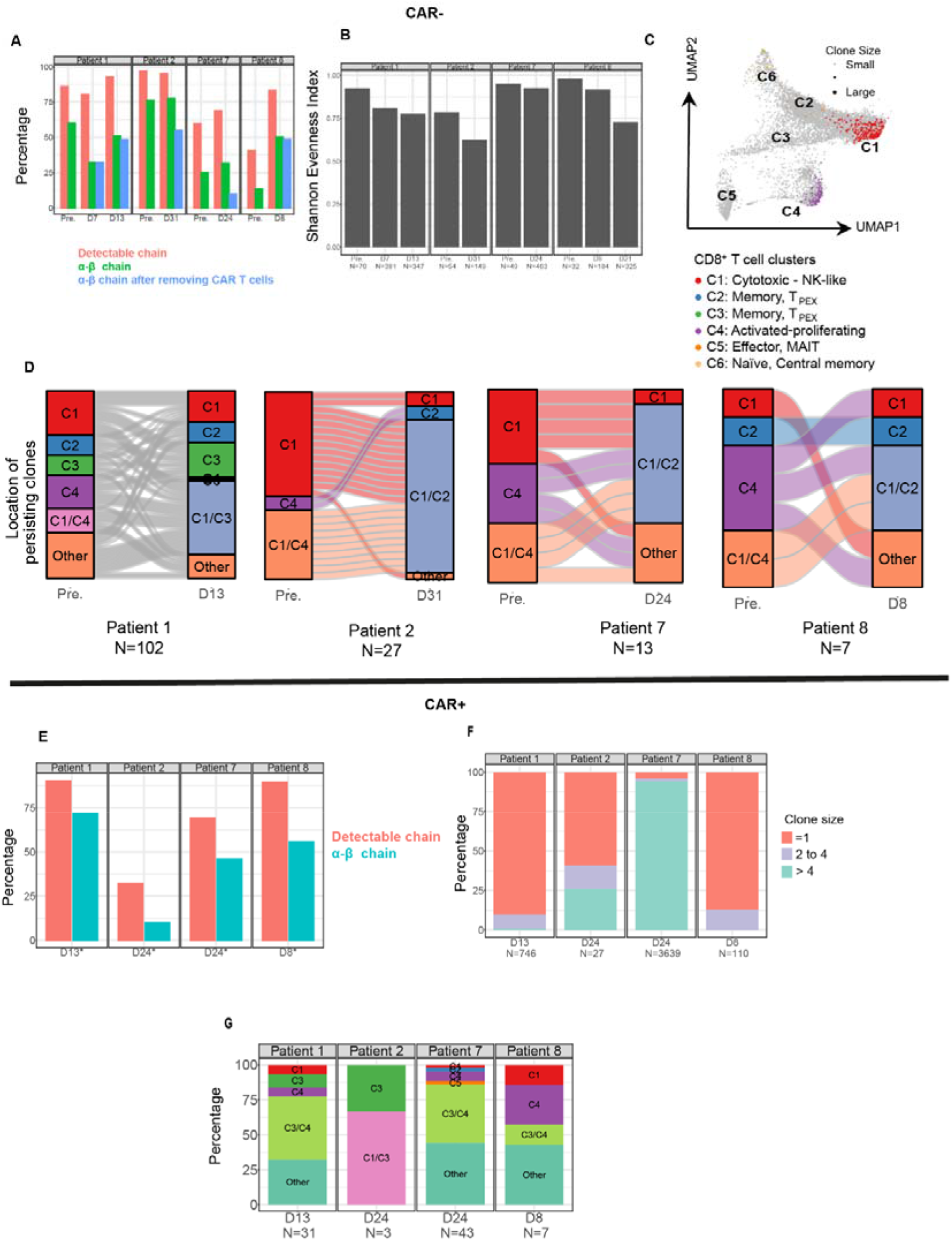
Clonal distribution of CAR^-^ and CAR^+^ CD8*^+^* T cells. **(A)** Percentage of cells in the CAR^-^ CD8^+^ T cells with a detectable α-β chain. **(B)** Shannon evenness values calculated from the TCR repertoire of CAR^-^ CD8^+^ T cells in each sample. **(C)** Distribution of expanded clones with size >1 identified in the CAR^-^CD8^+^ T cells in pre-infusion samples mapped on the UMAP plot, identifying the distribution on the CD8^+^ clusters. **(D)** Alluvial plots revealing the distribution of clones identified in CAR^-^ CD8^+^ T cells in pre- and post-infusion samples, and their cluster distribution based on gene expression profiles (C1 to C6). “Other” denotes clones observed in multiple phenotypes. Number of clones indicated by N. **(E)** Percentage of cells in the CAR^+^ CD8^+^ T cells with a detectable α-β chain. **(F)** Distribution of clones identified in the CAR^+^CD8^+^ T cells in post-infusion samples. **(G)** Distribution of clones identified in the CAR^+^CD8^+^ T cells in each cluster identified from scRNA-seq. Shown are post-infusion samples.

**Extended Data Fig 6.**
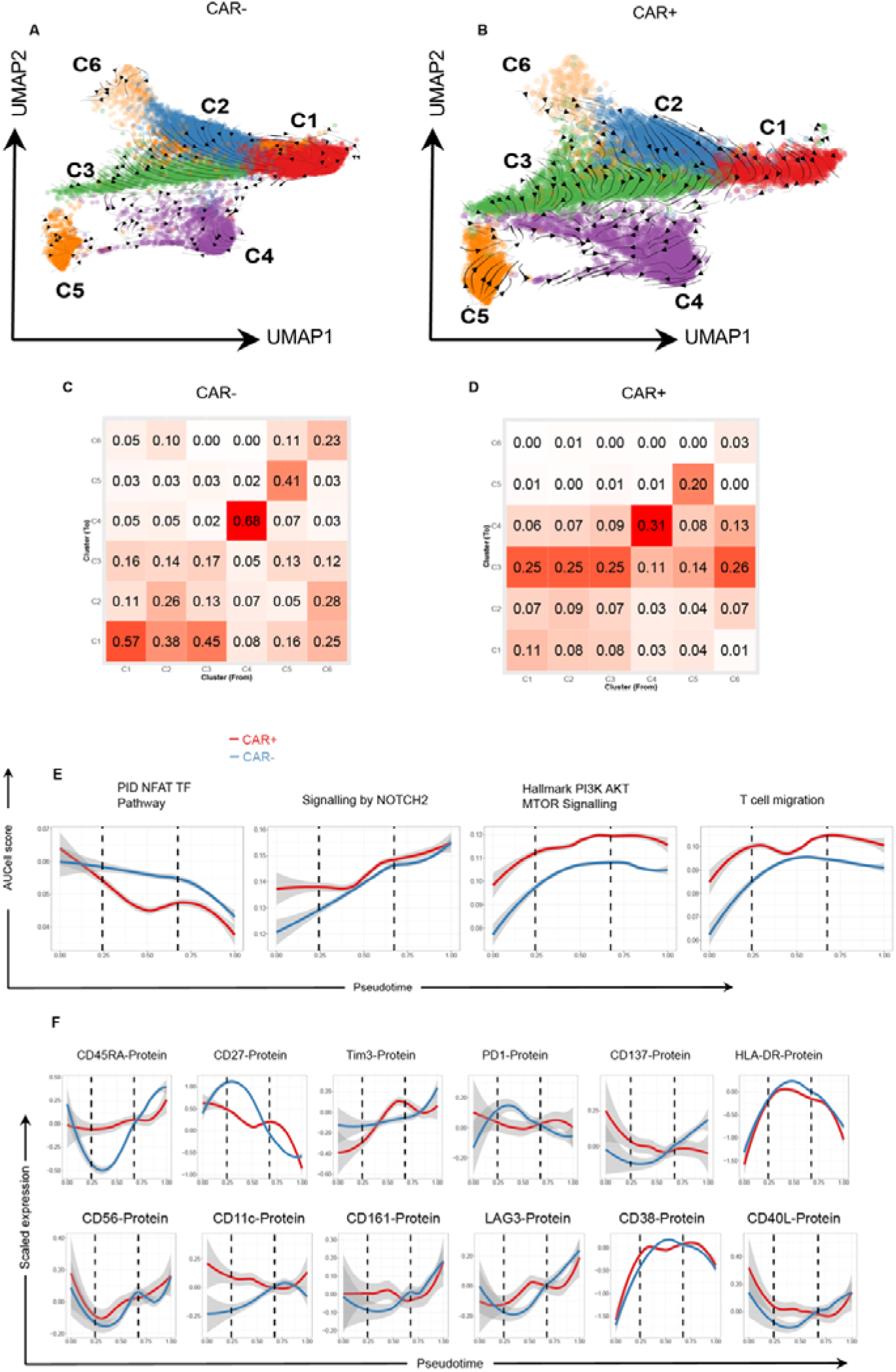
RNA velocity and cluster transition probabilities confirming the inferred differentiation trajectories. **A)** UMAP showing extrapolated future states (arrows) estimated by RNA velocity, applied to CAR^-^ CD8^+^ cells. **B)** UMAP showing extrapolated future states (arrows) estimated by RNA velocity, applied to all CAR CD8^+^ single cells. **C)** Heatmap showing probability of directed cluster transitions calculated using CellRank, averaged over all CAR^-^CD8^+^ T cells. **D)** Heatmap showing probability of directed cluster transitions calculated using CellRank, averaged over all CAR^+^ CD8^+^ T samples. **(E)** Loess curves for gene signature along estimated pseudotime trajectories. Scores are calculated usilng AUCell. (**F**) Loess curves for protein expression along estimated pseudotime trajectories.

**Extended Data Fig. 7.**
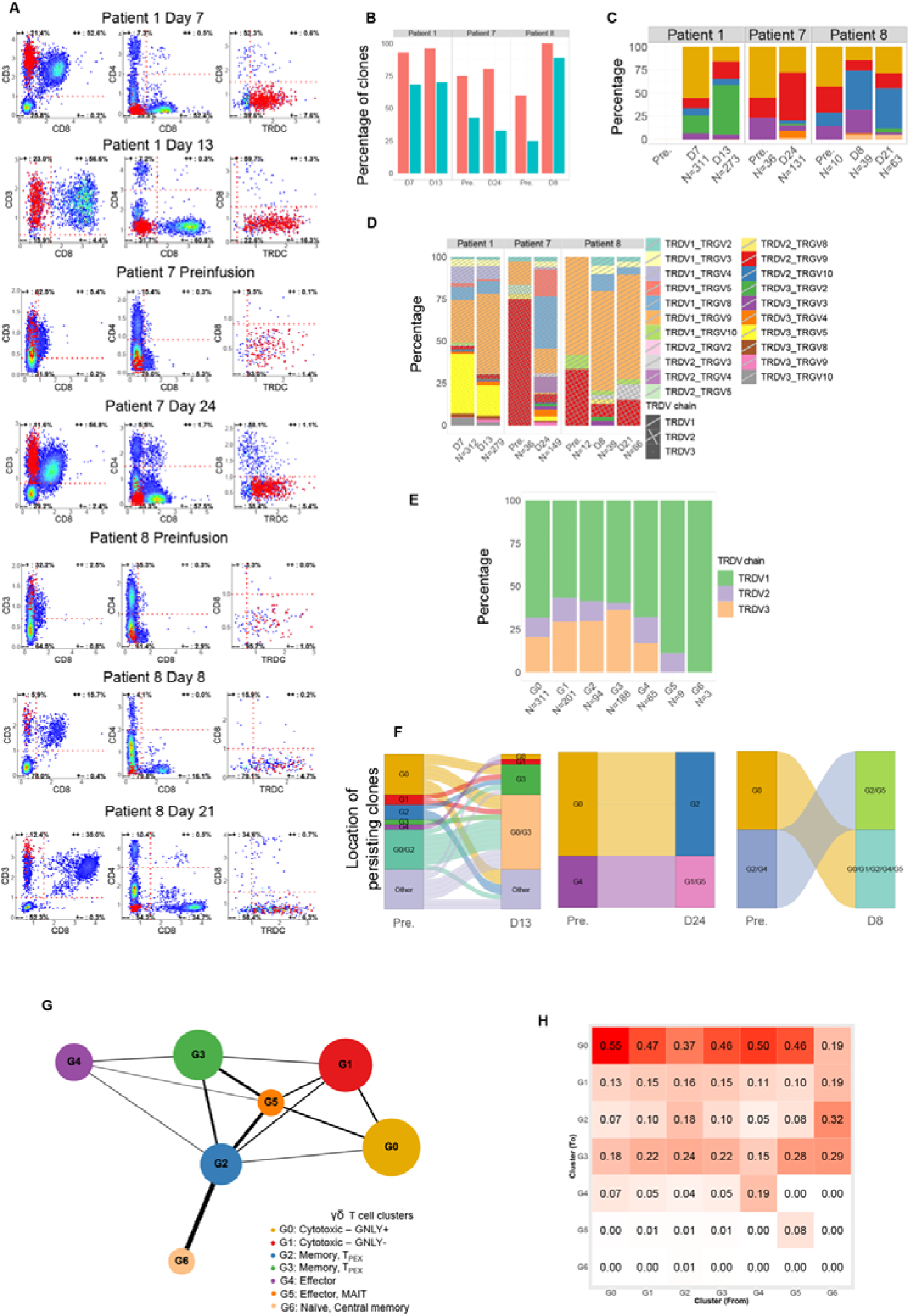
Identification of γδ T cells with clonal and differentiation trajectory analyses. **(A)** Scatter plots of markers used to define γδ T cells for each sample with γδ T cells. **(B)** Percentage of cells in the γδ T population with a detectable chain and detectable γ-δ chain. **(C)** Distribution of clones in each cluster. **(D)** Distribution of clones divided by gamma and delta chain, for each patient and sample time point. The number of clones is indicated by N. **(E)** Distribution of clones divided by delta chain gene usage, for each cluster. The number of clones is indicated by N. (**F**) Alluvial plots revealing the distribution of clones in pre- and post-infusion samples, and their cluster distribution based on gene expression profiles (G0 to C6). “Other” denotes clones observed in multiple phenotypes. Number of clones indicated by N. (**G**) PAGA graph revealing the cluster transitions weights (represented by line weights). **(H)** Heatmap showing probability of directed cluster transitions calculated using CellRank, averaged over all endogenous samples.

## SUPPLEMENTARY TABLES

**Table S1.** Patients’ clinical information.

| # | Sex | Disease | Primary refractory | Prior auto | Matched sibling allo conditioning | DLI | Prior blin | Total prior lines of therapy | Status at first dose CAR-T | Treated on-trial? | CAR19 T cell doses administered (lymphodepletion) | Best response^ (dose# achieved) | Duration of response (days) | CRS<sup>†</sup> | CAR T cell malignancy |
| --- | --- | --- | --- | --- | --- | --- | --- | --- | --- | --- | --- | --- | --- | --- | --- |
| P1 | F | B-ALL | No | No | MAC | No | Yes | 5 | Extramedullary | Yes | $1 \times 10^7$ cells/m<sup>2</sup> (Flu/Cy)<br>$5 \times 10^7$ cells/m<sup>2</sup> (Cy)<br>$5 \times 10^7$ cells/m<sup>2</sup> (Flu/Cy)<br>$1 \times 10^8$ cells/m<sup>2</sup> (Flu/Cy) | CR (1) | 154 | Gr 1 | No |
| P2 | M | DLBCL | Yes | BEAM | RIC | 3 | No | 8 | Stage IV | Yes | $1 \times 10^7$ cells/m<sup>2</sup> (Flu/Cy)<br>$5 \times 10^7$ cells/m<sup>2</sup> (Cy)<br>$1 \times 10^8$ cells/m<sup>2</sup> (Cy) | PR (1) | 335* | No | 3.2 months |
| P3 | M | B-ALL | No | No | RIC | No | No | 4 | MRD+ | Yes | $1 \times 10^7$ cells/m<sup>2</sup> (Flu/Cy)<br>$5 \times 10^7$ cells/m<sup>2</sup> (Cy)<br>$5 \times 10^7$ cells/m<sup>2</sup> (Flu/Cy)<br>$1 \times 10^8$ cells/m<sup>2</sup> (Flu/Cy) | CR (1) | 105 | No | No |
| P4 | M | B-ALL | No | No | MAC | No | Yes | 5 | CMR | No (prior seizures) | $1 \times 10^7$ cells/m<sup>2</sup> (Flu/Cy)<br>$5 \times 10^7$ cells/m<sup>2</sup> (Flu/Cy)<br>$1 \times 10^8$ cells/m<sup>2</sup> (Flu/Cy) | CR (1) | 230* | No | No |
| P5 | M | B-ALL | No, MRD+ | No | MAC | 3 | Yes | 5 | Blasts 70% | No (IFI) | $1 \times 10^7$ cells/m<sup>2</sup> (Flu/Cy)<br>$1 \times 10^7$ cells/m<sup>2</sup> (Flu/Cy) | CR (1) | 224* | Gr 3 | No |
| | | | | | | | | | | | $5 \times 10^7$<br>cells/m<sup>2</sup> (Flu/Cy)<br>$1 \times 10^8$<br>cells/m<sup>2</sup> (Cy) | | | | |
| A1 | M | DLB<br>CL | Yes | No | N/A | N/A | No | 3 | Stage<br>III | No | $1 \times 10^7$ cells/m<sup>2</sup><br>(Flu/Cy)<br>$5 \times 10^7$<br>cells/m<sup>2</sup> (Flu/Cy)<br>$38.6 \times 10^6$<br>cells/m<sup>2</sup> (Flu/Cy) | PR (2) | 31 | No | No |
| P7 | M | B-<br>ALL | No,<br>MRD+ | No | MAC | No | Yes | 4 | BM:<br>CMR<br>Occult<br>extram<br>edullary | Yes | $1 \times 10^7$<br>cells/m<sup>2</sup> (Flu/Cy) | CR (1) | 125* | Gr 2 | No |
| P8 | M | B-ALL | No | No | MAC | No | Yes | 5 | MRD<br>+ | Yes | $1 \times 10^7$<br>cells/m<sup>2</sup> (Flu/Cy) | CR (1) | 97* | No | 11.5<br>months |

**Table S2.** Single cell multi-omics data generated in each sample.

| Patient | Samples | Population | Days | Technologies |
| --- | --- | --- | --- | --- |
| P1 | Infusion product | CAR T-cells | NA | 10x only + Rhapsody/AbSeq + CyTOF |
|  | Circulating | CAR T-cells | 13 | AbSeq/10x/RageSeq + CyTOF |
|  | Pre-chemo | PBMC | -3 | CyTOF |
|  | Pre-infusion | PBMC | -1 | AbSeq/10x/RageSeq + CyTOF + Cytokines |
|  | Post-infusion | PBMC | 7 | AbSeq/10x/RageSeq + Cytokines |
|  | Post-infusion | PBMC | 13 | AbSeq/10x/RageSeq + CyTOF + Cytokines |
|  | Post-infusion | PBMC | 28 | CyTOF + Cytokines |
| P2 | Infusion product | CAR T-cells | NA | AbSeq/10x/Ragseq + CyTOF + TCR beta deep sequencing |
|  | Circulating | CAR T-cells | 24 | AbSeq/10x + CyTOF + RageSeq |
|  | Pre-chemo | PBMC | -3 | CyTOF |
|  | Pre-infusion | PBMC | -1 | AbSeq/10x/RageSeq + CyTOF + Cytokines |
|  | Post-infusion | PBMC | 24 | AbSeq/10x + CyTOF + Cytokines |
|  | Post-infusion | PBMC | 31 | AbSeq/10x/RageSeq + CyTOF |
| P3 | Infusion product | CAR T-cells | NA | CyTOF |
|  | Circulating | CAR T-cells | 14 | CyTOF |
|  | Circulating | CAR T-cells | 24 | CyTOF |
|  | Pre-chemo | PBMC | -3 | CyTOF |
|  | Pre-infusion | PBMC | -1 | CyTOF+ Cytokine |
|  | Post-infusion | PBMC | 14 | CyTOF+ Cytokine |
|  | Post-infusion | PBMC | 24 | CyTOF+ Cytokine (D28) |
| P4 | Infusion product | CAR T-cells | NA | CyTOF |
|  | Circulating | CAR T-cells | 16 | CyTOF |
|  | Circulating | CAR T-cells | 28 | CyTOF |
|  | Pre-infusion | PBMC | -1 | CyTOF+ Cytokine |
|  | Post-infusion | PBMC | 16 | CyTOF+ Cytokine |
|  | Post-infusion | PBMC | 28 | CyTOF+ Cytokine |
| P5 | Infusion product | CAR T-cells | NA | CyTOF |
|  | Circulating | CAR T-cells | 11 | CyTOF |
|  | Circulating | CAR T-cells | 29 | CyTOF |
|  | Pre-infusion | PBMC | -1 | CyTOF+ Cytokine |
|  | Post-infusion | PBMC | 11 | CyTOF+ Cytokine (D10) |
|  | Post-infusion | PBMC | 29 | CyTOF+ Cytokine |
| A1 | Infusion product | CAR T-cells | NA | CyTOF |
|  | Circulating | CAR T-cells | 17 | CyTOF |
|  | Circulating | CAR T-cells | 28 | CyTOF |
|  | Pre-infusion | PBMC | -1 | CyTOF+ Cytokine |
|  | Post-infusion | PBMC | 17 | CyTOF+ Cytokine |
|  | Post-infusion | PBMC | 28 | CyTOF+ Cytokine |
| P7 | Infusion product | CAR T-cells | NA | AbSeq/Rhapsody + CyTOF |
|  | Circulating | CAR T-cells | 24 | AbSeq/10x/RageSeq+ CyTOF |
|  | Pre-infusion | PBMC | -1 | AbSeq/10x/RageSeq + CyTOF + Cytokine |
|  | Post-infusion | PBMC | 24 | AbSeq/10x/RageSeq + CyTOF + Cytokine |
| P8 | Infusion product | CAR T-cells | NA | AbSeq/Rhapsody + CyTOF + TCR beta deep sequencing |
|  | Circulating | CAR T-cells | 8 | AbSeq/10x/RageSeq + CyTOF |
|  | Pre-infusion | PBMC | -1 | AbSeq/10x/RageSeq + CyTOF + Cytokine |
|  | Post-infusion | PBMC | 8 | AbSeq/10x/RageSeq + CyTOF + Cytokine |
|  | Post-infusion | PBMC | 21 | Abseq/10x |
| Healthy | NA | PBMC | NA | 10x only |

**Table S3.** Oligonucleotide-conjugated antibody (Abseq)

| Target | Clone | Barcode ID<br>(precommercial) | Catalogue<br>number |
| --- | --- | --- | --- |
| CD127 | HIL-7R-M21 | AHS0028 | 940012 |
| CD14 | M□P9 | AHS0037 | 940005 |
| CD161 | DX12 | AHS0002 | 940070 |
| CD183 | 1C6/CXCR3 | AHS0031 | 940030 |
| CD185<br>(CXCR5) | RF8B2 | AHS0039 | 940042 |
| CD19 | HIB19 | (v2a_Hs_0030) |  |
| CD194 | 1G1 | AHS0038 | 940047 |
| CD196 (CCR6) | 11A9 | AHS0034 | 940033 |
| CD197 (CCR7) | 150503 | (v2a_Hs_007) |  |
| CD25 | M-A251 | (v2a_Hs_0026) |  |
| CD27 | M-T271 | AHS0025 | 940018 |
| CD28 | CD28.2 | AHS0024 | 940017 |
| CD3 | SK7 | AHS0033 | 940000 |
| CD38 | HIT2 | AHS0022 | 940013 |
| CD4 | SK3 | AHS0032 | 940001 |
| CD45RA | HI100 | AHS0009 (v2a_Hs_0029) | 940011 |
| CD45RO | UCHL1 | AHS0036 | 940022 |
| CD8 | RPA-T8 | AHS0027 | 940003 |
| CD95 | DX2 | AHS0023 | 940037 |
| HLA-DR | G46-6 | (v2a_Hs_0035) |  |
| PD1 (CD279) | MIH4 | (v2a_Hs_0014) |  |
| CD10 | HI10a | AHS0051 | 940045 |
| CD11b | M1/70 | AHS0005 | 940008 |
| CD11c | B-ly6 | AHS0056 | 940024 |
| CD137 | 4B4-1 | AHS0003 | 940055 |
| CD141 | 1A4 | AHS0083 | 940079 |
| CD154 | TRAP1 | AHS0077 | 940053 |
| CD16 | 3G8 | AHS0053 | 940006 |
| CD20 | 2H7 | AHS0008 | 940016 |
| CD206 | 19.2 | AHS0072 | 940068 |
| CD21 | B-ly4 | AHS0074 | 940048 |
| CD274 (B7-H1) | MIH1 | AHS0004 | 940035 |
| CD40 | 5C3 | AHS0117 | 940049 |
| CD56 | NCAM16.2 | AHS0019 | 940007 |
| CD80 (B7-1) | L307.4 | AHS0046 |  |
| CD86 (B7-2) | 2331 (FUN-1) | AHS0057 | 940025 |
| IgD | IA6-2 | AHS0058 | 940026 |
| IgG | G18-145 | AHS0059 | 940027 |
| LAG-3 (CD223) | T47-530 | AHS0018 | 940080 |
| TIM-3 (CD366) | 7D3 | AHS0016 | 940066 |
| CD19 | SJ25C1 | AHS0030 | 940004 |
| CD25 | 2A3 | AHS0026 | 940009 |
| CD197 (CCR7) | 3D12 | AHS0007 | 940014 |
| PD1 (CD279) | EH12.1 | AHS0014 | 940015 |

**Table S4.** List of antibodies used in mass cytometry.

| Conjugated label | Specificity | Antibody clone | Manufacturer | Comment |
| --- | --- | --- | --- | --- |
| 104 | CD45 | HI30 | BD | CAR T product barcode |
| 110 | CD45 | HI30 | BD | Donor PBMC barcode |
| 113 | CD56 | REA196 | Miltenyi |  |
| 115 | CD8A | RPA-T8 | Biolegend |  |
| 139 | CD57 | HCD57 | Biolegend |  |
| 141 | CD49d | 9F10 | DVS/Fluidigm |  |
| 142 | CD19 | HIB19 | BD |  |
| 143 | CD45RA | HI100 | Biolegend |  |
| 144 | CD69 | FN50 | Biolegend |  |
| 145 | CD4 | RPA-T4 | BD |  |
| 146 | EOMES | WD1928 | Life Technologies | Intracellular |
| 147 | CD185 (CXCR5) | RF8B2 | BD |  |
| 148 | CD28 | CD28.2 | Biolegend | MMM |
| 149 | CD366 (TIM3) | 7D3 | BD | MMM |
| 150 | KLRG1 | SA231A2 | Biolegend | MMM |
| 151 | CD39 | A1 | Biolegend | MMM |
| 152 | CD45RO | UCHL1 | BD |  |
| 153 | CD62L | DREG-56 | Biolegend |  |
| 154 | CD196 (CCR6) | REA190 | Miltenyi |  |
| 155 | CD137 (4-1BB) | 4B4-1 | BD | MMM |
| 156 | CD279 (PD1) | EH12.2H7 | Biolegend | MMM |
| 158 | CD194 (CCR4) | L291H4 | Biolegend |  |
| 159 | CD197 (CCR7) | 150503 | R&D Systems |  |
| 160 | CD223 (Lag3) | 17B4 | Nobus Bio | MMM |
| 161 | CD122 (IL-2RB) | TU27 | Biolegend | MMM |
| 162 | Foxp3 | PCH101 | eBioscience | Intracellular |
| 163 | CD183 (CXCR3) | REA232 | Miltenyi |  |
| 164 | CD274 (PDL1) | MIH1 | BD | MMM |
| 165 | CD16 | 3G8 | DVS/Fluidigm |  |
| 166 | TIGIT | MBSA43 | RIZO | MMM |
| 167 | CAR T | 136.20.1 | MDACC |  |
| 168 | Ki67 | B56 | BD | Intracellular |
| 169 | CD25 | M-A251 | Biolegend |  |
| 170 | CD3 | UCHT1 | BD |  |
| 171 | Granzyme B | GB11 | Acris | Intracellular |
| 172 | CD38 | HIT2 | DVS/Fluidigm |  |
| 173 | Integrin b7 | FIB504 | BD |  |
| 174 | HLA-DR | G46-6 | BD |  |
| 175 | Perforin | B-D48 | DVS/Fluidigm | Intracellular |
| 176 | CD127 | A019D5 | Biolegend |  |
| 209 | T-bet | 4B10 | BD | Intracellular |

The following tables have been uploaded as csv files due to their large size.

**Table S5 Differentially expressed genes, see excel spreadsheet containing all the results.**

**Table S6 Gene set enrichment analysis of differentially expressed genes.**

**Table S7 TCR sequences interrogated with VDJdb for known antigen specificity.**

**Table S8 List of curated gene signatures utilised to calculate gene modules’ scores.**

